# A First-Visit Clinical Score to Distinguish Immunoglobulin Light-Chain from Wild-Type Transthyretin Cardiac Amyloidosis: Derivation and Internal Validation

**DOI:** 10.64898/2026.09.10.26362801

**Authors:** Yosuke Matsumoto, Yasuhiro Izumiya, Yawara Kawano, Naoto Kuyama, Kei Morikawa, Akihisa Tabira, Masahiro Yamamoto, Kyoko Hirakawa, Masanobu Ishii, Shinsuke Hanatani, Yasushi Matsuzawa, Hiroki Usuku, Eiichiro Yamamoto, Hirofumi Soejima, Kenichi Tsujita

## Abstract

**Background:** Delayed diagnosis of immunoglobulin light-chain cardiac amyloidosis (AL-CA) worsens prognosis, partly because AL-CA and wild-type transthyretin cardiac amyloidosis (ATTRwt-CA) are difficult to distinguish at the initial cardiology visit. We developed and validated a practical score to triage AL-CA from ATTRwt-CA.

**Methods:** We retrospectively analyzed 541 consecutive patients with cardiac amyloidosis (91 AL-CA, 450 ATTRwt-CA) at our institution’s amyloidosis center, randomly divided into derivation (n=378) and validation (n=163) cohorts. Candidate first-visit variables underwent LASSO selection after multiple imputation, with estimates pooled by Rubin’s rules. Performance was assessed by bootstrap validation, calibration, and decision curve analysis. The model was converted into a simplified point-based score.

**Results:** Seven variables were retained: age, absence of atrial fibrillation, absence of carpal tunnel syndrome, low serum albumin, proteinuria (≥1+), low voltage, and thinner left ventricular posterior wall thickness. Areas under the curve (AUC) were 0.97 in the derivation cohort and 0.99 in the validation cohort; the bootstrap optimism-corrected AUC was 0.96. The model provided net benefit across threshold probabilities of 0.01– 0.50. Risk tiering categorized the validation cohort into low (0–5 points; AL-CA 0%), intermediate (6–9 points; 26%), and high (10–13 points; 95%) probability groups. Discrimination remained high for AL-CA versus monoclonal protein–positive ATTRwt-CA at a cutoff of ≥8 points (AUC 0.90). Adding this score to monoclonal protein testing significantly improved diagnostic performance compared with monoclonal protein testing alone.

**Conclusions:** This simple first-visit score may help cardiologists rapidly triage suspected AL-CA and prioritize urgent hematology referral and subtype-specific confirmatory testing.

**Clinical Perspective:** *What Is New?:*

- A 7-variable score using age, history of atrial fibrillation and carpal tunnel syndrome, serum albumin, dipstick proteinuria, low QRS voltage, and left ventricular posterior wall thickness distinguished AL-CA from ATTRwt-CA using information available at the first cardiology visit.
- The score retained high discrimination in the challenging subgroup comprising patients with AL-CA or monoclonal protein–positive ATTRwt-CA.

*What Are the Clinical Implications?:*

- The score may complement—not replace—standard monoclonal protein testing and definitive amyloid typing by helping prioritize the urgency of hematology assessment and subtype-specific confirmatory evaluation.

## Introduction

Cardiac amyloidosis (CA) is a secondary cardiomyopathy caused by extracellular deposition of amyloid fibrils within the myocardium, and represents an increasingly recognized and treatable cause of heart failure, arrhythmias, and progressive cardiomyopathy (1–2). The two predominant subtypes in clinical practice are immunoglobulin light-chain (AL) amyloidosis and wild-type transthyretin (ATTRwt) amyloidosis, both of which have seen rising incidence with population aging and improved diagnostic awareness (3–4). These subtypes require fundamentally distinct therapeutic strategies: plasma cell-directed chemotherapy—including daratumumab-based regimens—for light-chain cardiac amyloidosis (AL-CA), and TTR stabilizers or silencers for wild-type transthyretin cardiac amyloidosis (ATTRwt-CA) (5–7). Among these, AL-CA carries a particularly aggressive course, and delayed diagnosis is an independent determinant of mortality (8–9); therefore, early diagnosis of AL-CA is paramount to enabling timely therapeutic intervention and improving survival (10–11).

Despite growing clinical awareness, more than half of patients require 6 months or longer to receive a definitive diagnosis of AL-CA even after cardiology engagement (12), with direct consequences for disease stage at treatment initiation and survival (9). Initial symptoms are nonspecific and overlap with hypertrophic cardiomyopathy and heart failure with preserved ejection fraction (HFpEF) (3), and the high prevalence of ATTRwt-CA among elderly patients with HFpEF further complicates subtype differentiation at first presentation (13). Although current guidelines recommend a stepwise pathway of clinical, electrocardiographic, echocardiographic, laboratory, and extracardiac red flags followed by monoclonal protein screening and multimodality imaging (14–15), general cardiologists still need to recognize first-visit findings that indicate a high likelihood of AL-CA and warrant urgent hematological evaluation and subtype-specific testing.

The aim of this study was to develop and validate a practical prediction model capable of distinguishing AL-CA from ATTRwt-CA using only variables available at the initial outpatient visit. This score was designed for patients in whom CA is already suspected and was intended to support subtype identification rather than the primary diagnosis of CA itself.

## Methods

### Study Design and Participants

The retrospective, single-center study included 542 consecutive patients newly diagnosed with CA, specifically ATTRwt-CA and AL-CA, at the Amyloidosis Center of Kumamoto University Hospital between June 2002 and September 2025. The study size was determined by the number of consecutive eligible patients available during the prespecified study period; no a priori sample size calculation was performed because this was a retrospective analysis of an existing cohort. One patient with coexisting ATTRwt and AL amyloidosis was excluded from the analysis (16).

The study protocol complied with the principles of the Declaration of Helsinki and was approved by the Ethics Committee of Kumamoto University (Approval No. 1590). Given the retrospective nature of the study, the requirement for written informed consent was waived. Information regarding the study protocol was disclosed at Kumamoto University Hospital and on the departmental website (https://kumadai-junnai.com/archives/clinical), providing patients with the opportunity to opt out. The data are not publicly available because they contain information that could compromise participant privacy; deidentified data may be available from the corresponding author upon reasonable request and institutional approval.

AL-CA was defined by histologically proven systemic AL amyloidosis on extracardiac biopsy in combination with typical findings on cardiac magnetic resonance imaging or transthoracic echocardiography or by endomyocardial biopsy demonstrating AL deposits. ATTRwt-CA was diagnosed by typical echocardiographic or cardiac magnetic resonance findings of left ventricular thickening combined with Grade 2 or 3 myocardial uptake on 99mTc-pyrophosphate scintigraphy or by biopsy. All biopsy samples were confirmed by Congo red staining and subtyped using mass spectrometry or immunohistochemistry. The absence of pathogenic transthyretin gene mutations on sequencing confirmed the diagnosis of ATTRwt amyloidosis (14). Plasma cell disorders were reviewed in all patients as part of subtype adjudication. Serum free light-chain assay and serum and urine immunofixation electrophoresis were assessed according to the contemporary diagnostic algorithm (14). In all patients in whom monoclonal protein was detected, AL amyloid deposition was assessed by histological examination. Patients were classified as ATTRwt-CA only when TTR amyloid deposition was detected.

### Data Collection

Clinical and diagnostic data were obtained at the time of the initial visit. Predictor variables were extracted from the initial-visit clinical records, 12-lead electrocardiography, echocardiography, and laboratory data using predefined definitions before model fitting. A history of atrial fibrillation (AF) was defined as a documented diagnosis of paroxysmal, persistent or permanent AF before or at the initial visit, based on prior electrocardiograms, Holter recordings, referral documents, or medical records. Atrial flutter was not included in this definition. A history of carpal tunnel syndrome (CTS) was defined as a documented prior physician diagnosis of CTS or a recorded history of carpal tunnel release surgery. Serum albumin was measured by standard laboratory methods and analyzed as a continuous variable (g/dL). Proteinuria was assessed by automated urine dipstick testing and defined as ≥1 + (recorded as negative/trace/1+/2+/3+/4+). Low voltage on electrocardiography was defined as QRS amplitude <5 mm in the limb leads and/or <10 mm in the precordial leads. Left ventricular ejection fraction was calculated using the modified Simpson’s method. An abnormal **κ/λ** ratio was defined as <0.26 or >1.65 (17).

### Prediction Model Development

The model was developed in accordance with the Transparent Reporting of a multivariable prediction model for Individual Prognosis Or Diagnosis (TRIPOD) guidelines (Supplemental Table 1) (18).

The prediction score was developed in a derivation cohort comprising 378 randomly selected patients (70%) and subsequently validated in the remaining 163 patients (30%) without outcome stratification. Variables with high proportions of missing data, namely PR interval (46%) and E/A ratio (41%), were excluded from model development on the grounds of practicality in the derivation cohort. These missing data were largely attributable to AF at the time of assessment. Given the high frequency of AF in patients with ATTRwt-CA (19), it was considered appropriate to exclude these variables in order to prioritize practical applicability. As the results of serum free light-chain and immunofixation electrophoresis were not available on the day of the initial visit, these variables were excluded. Prior to multivariable model building, collinearity among candidate predictors was formally evaluated using the variance inflation factor (VIF) in the derivation cohort across the imputed datasets. Variables exhibiting substantial multicollinearity, defined a priori as a VIF threshold >10, were excluded before applying the Least Absolute Shrinkage and Selection Operator (LASSO) regression.

Other missing data were handled using multiple imputation by chained equations (MICE), generating 20 datasets after random allocation to the derivation and internal validation cohorts. Predictive mean matching was used for continuous variables and logistic regression for binary variables. The imputation model included all candidate predictors and the outcome as a predictor (20).

In the derivation cohort, LASSO logistic regression was performed on each imputed dataset. LASSO selected the shrinkage parameter using the 1-SE rule in 10-fold cross- validation. Variables selected in all 20 imputed derivation datasets were retained in the final prediction model (18). Finally, the selected variables were entered into logistic regression analysis for each imputed dataset, and the results were pooled using Rubin’s rules (21).

### Internal Validation

Model discrimination was evaluated using the area under the receiver operating characteristic curve (AUC), and performance was assessed using the Brier score. To account for overfitting, internal validation was performed using 1,000 bootstrap resamples. The optimism-corrected AUC was calculated by subtracting the average optimism from the original AUC. Calibration was visually assessed using LOESS smoothed calibration plots and quantified by the calibration slope and calibration-in- the-large (CITL). To adjust for baseline differences between cohorts, recalibration-in- the-large was performed in the validation cohort by updating the intercept to align predicted probabilities with observed prevalence. The clinical utility of the diagnostic model was evaluated using decision curve analysis (DCA), which quantified net benefit across a range of threshold probabilities (22). In this study, the threshold probability represented the predicted probability above which additional diagnostic evaluation for suspected AL-CA would be initiated. Given the substantial clinical harm associated with false negative results due to delayed diagnosis of AL-CA, threshold probabilities ranging from 0.01 to 0.50 were prespecified. Net benefit of the model was compared with treat-all and treat-none strategies in both the derivation and validation cohorts.

### Simplified Score Development

A simplified, practical scoring system was developed using variables from the multivariable model. Continuous variables were dichotomized at optimal cutoff values defined by the maximum sum of sensitivity and specificity on receiver operating characteristic analysis in the derivation cohort. Weighted scores were calculated by dividing each regression coefficient by the smallest absolute coefficient, while considering clinical value. Diagnostic performance of the simplified score was assessed, and patients were stratified into low, intermediate, and high probability groups based on observed event rates and clinical interpretability. Finally, in a combined model that integrated the simplified score (≥8) with the abnormal serum free light-chain κ/λ ratio (FLC model) and positive findings on serum or urine immunofixation electrophoresis (immunofixation model), we evaluated the AUC, ΔAUC, NRI, and IDI to verify the additional predictive value of the simplified score. Furthermore, as a sensitivity analysis, we similarly evaluated the diagnostic performance of the simplified score in cohorts limited to patients with AL-CA and patients with ATTRwt-CA who tested positive for monoclonal proteins.

All available patients during the study period were included. Continuous variables are presented as mean ± standard deviation or median with interquartile range (non-normal distribution), and compared using Student’s t-test or Mann–Whitney U test, respectively. Categorical variables were compared using the chi-square test or Fisher exact test, as appropriate. Two-sided p<0.05 was considered statistically significant. All statistical analyses were performed using R version 4.5.2 (R Foundation for Statistical Computing, Vienna, Austria).

## Results

### Baseline Characteristics

A total of 542 patients with CA were enrolled; after excluding one case, 541 patients were analyzed. The derivation cohort consisted of 63 patients with AL-CA and 315 with ATTRwt-CA, while the validation cohort included 28 with AL-CA and 135 with ATTRwt-CA (Figure 1). Baseline characteristics of the derivation cohort are presented in Table 1.

**Figure 1.**
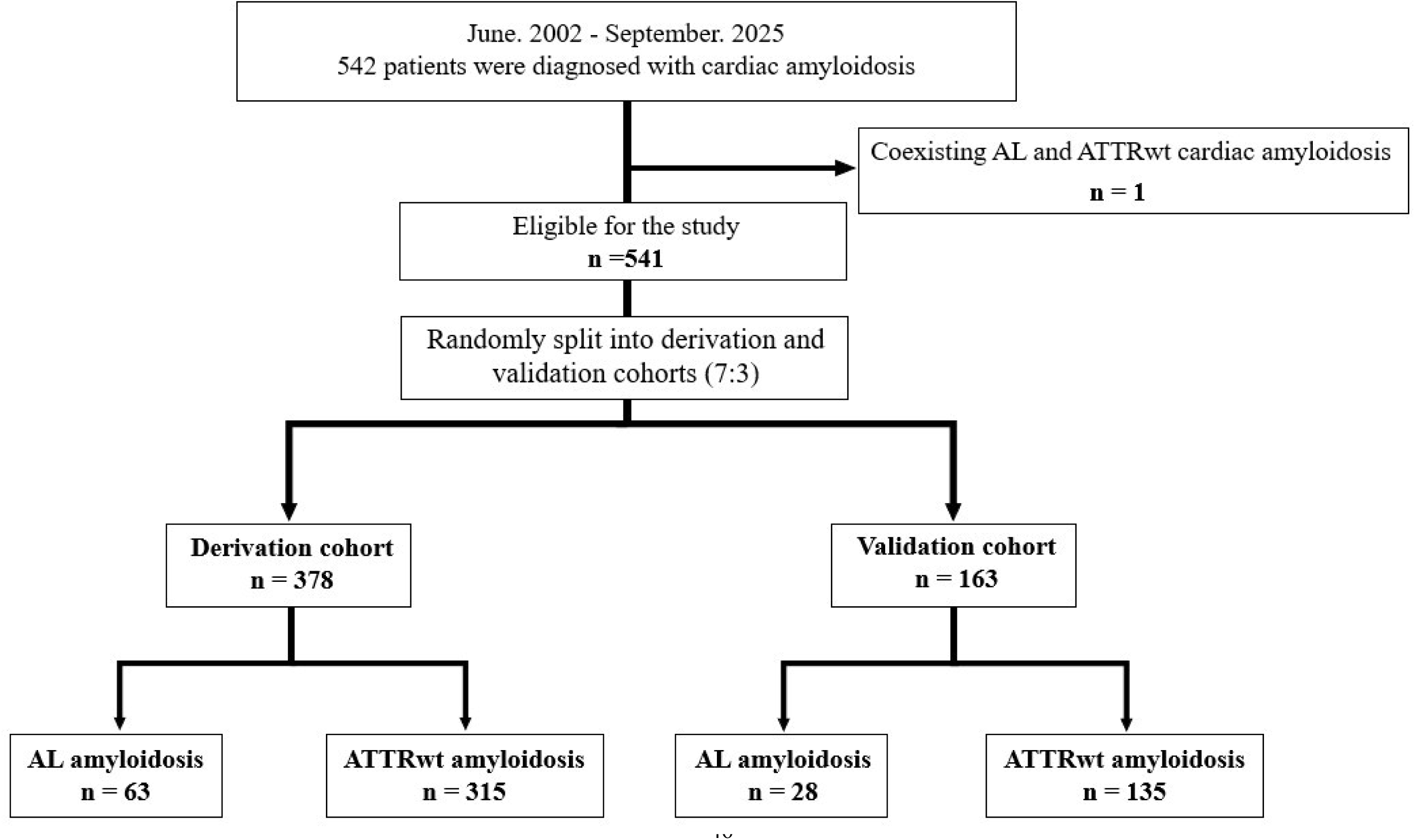
Flow chart of the study population. AL, immunoglobulin light chain; ATTR, transthyretin.

**Table 1.** Patient baseline characteristics of the derivation cohort.

|  | **Overall** | **AL** | **ATTRwt** | **missing** | **p-value** |
| --- | --- | --- | --- | --- | --- |
| Total | 378 | 63 | 315 | 0 |  |
| **Age (years)** | **76.3 ± 8.09** | **68.5 ± 9.90** | **77.8 ± 6.67** | **0** | **<0.001** |
| **Male** | **310 (82)** | **41 (65)** | **269 (85)** | **0** | **<0.001** |
| **Body mass index (kg/m<sup>2</sup>)** | **23.2 ± 3.35** | **22.3 ± 3.90** | **23.4 ± 3.21** | **0** | **0.016** |
| Comorbidity |  |  |  |  |  |
| **Hypertension** | **212 (56)** | **27 (43)** | **185 (59)** | **0** | **0.020** |
| Diabetes mellitus | 81 (21) | 9 (14) | 72 (23) | 0 | 0.130 |
| Dyslipidemia | 125 (33) | 22 (35) | 103 (33) | 0 | 0.732 |
| Chronic kidney disease | 274 (73) | 44 (70) | 230 (73) | 0 | 0.607 |
| Hemodialysis | 1 (1) | 1 (2) | 0 (0) | 0 | 0.167 |
| **Atrial fibrillation** | **174 (46)** | **6 (10)** | **168 (53)** | **0** | **<0.001** |
| Ischemic heart disease | 30 (8) | 6 (10) | 24 (8) | 0 | 0.610 |
| **Carpal tunnel syndrome** | **174 (46)** | **5 (8)** | **169 (54)** | **0** | **<0.001** |
| **Spinal canal stenosis** | **101 (26)** | **4 (7)** | **97 (30)** | **0** | **<0.001** |
| Permanent pacemaker | 33 (9) | 2 (3) | 31 (10) | 0 | 0.087 |
| Implantable cardioverter defibrillator | 12 (3) | 2 (3) | 10 (3) | 0 | 1.00 |
| **Post radiofrequency catheter ablation** | **43 (11)** | **1 (2)** | **42 (13)** | **0** | **0.004** |
| Post PCI / CABG | 35 (9) | 3 (5) | 32 (10) | 0 | 0.098 |
| History of heart failure | 208 (55) | 33 (52) | 175 (56) | 0 | 0.644 |
| **Hospitalization of heart failure** | **113 (30)** | **10 (16)** | **103 (33)** | **0** | **0.008** |
| **NYHA functional class** |  |  |  | **0** | **0.033** |
| I | 87 (23) | 23 (37) | 64 (20) |  |  |
| II | 176 (47) | 27 (43) | 149 (47) |  |  |
| III | 111 (29) | 12 (19) | 99 (31) |  |  |
| IV | 4 (1) | 1 (2) | 3 (1) |  |  |
| **NYHA II or more** | **291 (77)** | **40 (64)** | **251 (80)** | **0** | **0.005** |
| **Monoclonal gammopathy** | **133 (35)** | **63 (100)** | **70 (22)** | **0** | **<0.001** |
| **Serum free $\kappa$ light chain (mg/L)** | **30.5 (20.5–48.8)** | **34.1 (14.3–131.0)** | **30.4 (21.9–46.2)** | **49** | **<0.001** |
| **Serum free $\lambda$ light chain (mg/L)** | **24.9 (18.2–38.4)** | **146.5 (39.2–347.0)** | **23.5 (17.7–31.1)** | **49** | **<0.001** |
| **Abnormal $\kappa / \lambda$ ratio** | **101 (31)** | **42 (70)** | **59 (22)** | **49** | **<0.001** |
| **Positive serum and / or urine immunofixation** | **76 (21)** | **54 (87)** | **22 (7)** | **9** | **<0.001** |
| Electrocardiographic parameters |  |  |  |  |  |
| **PR duration (ms)** | **205.0 ± 48.4** | **175.3 ± 30.9** | **213.6 ± 49.2** | **117** | **<0.001** |
| **QRS duration (ms)** | **109.3 ± 26.5** | **97.5 ± 26.1** | **111.9 ± 25.9** | **34** | **<0.001** |
| LBBB | 34 (10) | 3 (9) | 31 (11) | 34 | 0.140 |
| RBBB | 62 (18) | 8 (13) | 54 (19) | 34 | 0.242 |
| **QTc (ms)** | **452.8 ± 31.2** | **438.8± 36.3** | **455.9 ± 29.1** | **34** | **<0.001** |
| **Low QRS voltage** | **64 (17)** | **29 (45)** | **35 (11)** | **9** | **<0.001** |
| Pseudo-infarct pattern | 190 (52) | 37 (60) | 153 (50) | 9 | 0.164 |
| Echocardiographic parameters |  |  |  |  |  |
| **IVST (mm)** | **15.0 ± 2.88** | **12.7 ± 2.65** | **15.4 ± 2.70** | **0** | **<0.001** |
| **LVPWT (mm)** | **15.0 ± 3.02** | **12.4 ± 2.23** | **15.5 ± 2.88** | **0** | **<0.001** |
| **LVEF (%)** | **52.4 ± 10.1** | **58.0 ± 8.21** | **51.3 ± 10.0** | **0** | **<0.001** |
| LVEDD (mm) | 41.2 ± 5.90 | 40.5 ± 6.22 | 41.3 ± 5.84 | 0 | 0.294 |
| **LVESD (mm)** | **30.8 ± 6.32** | **26.8 ± 5.68** | **31.6 ± 6.15** | **0** | **<0.001** |
| **LVMI (g/m<sup>2</sup>)** | **155.0 ± 44.8** | **117.3 ± 37.8** | **162.6 ± 42.4** | **0** | **<0.001** |
| **Left atrial diameter (mm)** | **41.0 ± 6.99** | **36.8 ± 7.91** | **42.0 ± 6.44** | **0** | **<0.001** |
| Mitral deceleration time (ms) | 188.2 ± 64.6 | 196.1 ± 60.4 | 186.6 ± 65.5 | 0 | 0.293 |
| E/A | 1.61 ± 1.20 | 1.36 ± 1.05 | 1.67 ± 1.23 | 114 | 0.061 |
| **E/e'** | **19.5 ± 7.25** | **17.0 ± 6.79** | **20.0 ± 7.26** | **0** | **0.003** |
| Pericardial effusion | 161 (43) | 28 (44) | 133 (42) | 0 | 0.745 |
| Pleural effusion | 89 (24) | 20 (32) | 69 (22) | 0 | 0.093 |
| Moderate mitral regurgitation or more | 41 (11) | 5 (8) | 36 (11) | 0 | 0.416 |
| Moderate aortic stenosis or more | 28 (7) | 1 (2) | 27 (9) | 0 | 0.063 |
| Laboratory values |  |  |  |  |  |
| BNP (pg/mL) | 204.7 (111.5–351.9) | 144.6 (72.5–374.2) | 213.9 (118.7–349.9) | 3 | 0.213 |
| hs-troponin T (ng/mL) | 0.048 (0.033–0.075) | 0.040 (0.021–0.071) | 0.048 (0.034–0.075) | 3 | 0.080 |
| Creatinine (mg/dL) | 1.15 ± 0.52 | 1.15 ± 0.85 | 1.15 ± 0.43 | 0 | 1.00 |
| **eGFR (mL/min/1.73 m²)** | **51.7 ± 16.6** | **56.7 ± 22.5** | **50.7 ± 15.0** | **0** | **0.047** |
| **Hb (g/dL)** | **13.4 ± 1.80** | **12.8 ± 2.17** | **13.5 ± 1.71** | **0** | **0.016** |
| **Plt (×10³/μL)** | **190.9 ± 67.4** | **242.9 ± 106.3** | **180.6 ± 50.7** | **0** | **<0.001** |
| **TP (g/dL)** | **6.87 ± 0.83** | **5.95 ± 1.21** | **7.05 ± 0.58** | **0** | **<0.001** |
| **Alb (g/dL)** | **3.86 ± 0.61** | **3.14 ± 0.95** | **4.00 ± 0.39** | **0** | **<0.001** |
| Na (mmol/L) | 139.8 ± 2.78 | 139.7 ± 2.96 | 139.8 ± 2.75 | 0 | 0.656 |
| K (mmol/L) | 4.41 ± 0.44 | 4.40 ± 0.41 | 4.41 ± 0.45 | 0 | 0.818 |
| Cl (mmol/L) | 104.7 ± 3.33 | 105.0 ± 3.53 | 104.7 ± 3.15 | 0 | 0.440 |
| **PT (%)** | **80.6 ± 29.1** | **95.0 ± 23.2** | **78.2 ± 29.2** | **31** | **<0.001** |
| APTT (s) | 33.2 ± 7.59 | 31.8 ± 7.01 | 33.3 ± 7.28 | 31 | 0.108 |
| **Qualitative urine protein test** |  |  |  | **27** | **<0.001** |
| Negative | 229 (65) | 9 (15) | 220 (76) |  |  |
| Trace | 39 (10) | 5 (8) | 34 (12) |  |  |
| 1+ | 31 (9) | 9 (15) | 22 (8) |  |  |
| 2+ | 24 (7) | 13 (21) | 11 (4) |  |  |
| 3+ | 17 (5) | 15 (24) | 2 (1) |  |  |
| 4+ | 11 (3) | 11 (18) | 0 (0) |  |  |
| **Proteinuria** | **83 (24)** | **48 (77)** | **35 (12)** | **27** | **<0.001** |

Values are mean (SD), n / N (%), or median (IQR).
Abbreviations; AL = light-chain amyloidosis; ATTRwt = wild-type transthyretin amyloidosis; PCI = percutaneous coronary intervention; CABG = coronary artery bypass grafting; IVST = Interventricular septal thickness; LVPWT = Left ventricular posterior wall thickness; LVEF = Left ventricular ejection fraction; LVEDD = Left ventricular end-diastolic diameter; LVESD = Left ventricular end-systolic diameter; LVMI = Left ventricular mass index.

Electrocardiographic findings showed shorter PR and QRS durations, and a higher frequency of low voltage in the AL-CA group. Echocardiographically, AL-CA was characterized by thinner interventricular septal thickness and left ventricular posterior wall thickness (LVPWT), and by preserved left ventricular ejection fraction. Laboratory data revealed lower hemoglobin, total protein, and serum albumin, but higher eGFR in the AL-CA group. Proteinuria was significantly more prevalent in patients with AL-CA. Baseline characteristics for the validation cohort are provided in Supplemental Table 2. Among patients with ATTRwt-CA, monoclonal protein was detected in 70 patients in the derivation cohort and in 27 patients in the validation cohort.

### Prediction Model Development and Discriminative Performance

Across the 20 imputed datasets, LASSO regression consistently selected age, history of AF, history of CTS, serum albumin, proteinuria, low voltage and LVPWT, confirming selection stability (Supplemental Table 3). These seven variables were significantly associated with the diagnosis of AL-CA (Table 2). The predictive model demonstrated excellent discrimination, with an AUC of 0.97 (95% CI: 0.95–0.99) in the derivation cohort and 0.99 (95% CI: 0.97–0.99) in the internal validation cohort (Figure 2A, 2B). Bootstrap internal validation showed an optimism-corrected AUC of 0.96 (95% CI: 0.94–0.99).

**Table 2.** diagnostic model variables from the pooled multivariable logistic regression analysis and simplified score variables.

| **Panel A. Diagnostic model** |  |  |  |  |
| --- | --- | --- | --- | --- |
| **Variable** | **Regression coefficient** | **Odds ratio** | **95% CI** | **p-value** |
| Age (per 1 year increase) | -0.18 | 0.84 | 0.77–0.91 | <0.001 |
| Atrial fibrillation | -2.24 | 0.11 | 0.03–0.41 | 0.001 |
| Carpal tunnel syndrome | -3.16 | 0.04 | 0.01–0.20 | <0.001 |
| Alb (per 1 g/dL increase) | -1.48 | 0.23 | 0.08–0.63 | 0.005 |
| Proteinuria | 2.01 | 7.48 | 2.26–24.70 | 0.001 |
| Low QRS voltage | 1.66 | 5.24 | 1.56–17.63 | 0.007 |
| LVPWT (per 1 mm increase) | -0.35 | 0.70 | 0.56–0.88 | 0.003 |
| Intercept | 22.6 |  |  | <0.001 |

| Panel B. Simplified score |  |  |  |  |  |
| --- | --- | --- | --- | --- | --- |
| Variable | Regression coefficient | Odds ratio | 95% CI | p-value | Simplified model |
| Age (years) <74 | 2.00 | 7.38 | 2.46–22.2 | <0.001 | 2 |
| No Atrial fibrillation | 2.13 | 8.44 | 2.41–29.6 | 0.001 | 2 |
| No Carpal tunnel syndrome | 2.80 | 16.4 | 3.84–70.3 | <0.001 | 3 |
| Alb (g/dL) <3.6 | 1.27 | 3.56 | 1.16–11.0 | 0.027 | 1 |
| Proteinuria | 2.41 | 11.1 | 3.72–33.4 | <0.001 | 2 |
| Low QRS voltage | 1.64 | 5.17 | 1.60–16.7 | 0.006 | 1 |
| LVPWT (mm) <14.2 | 2.07 | 7.91 | 2.48–25.2 | <0.001 | 2 |

Abbreviations; Alb = albumin; LVPWT = left ventricular posterior wall thickness; CI = confidence interval.

**Figure 2.**
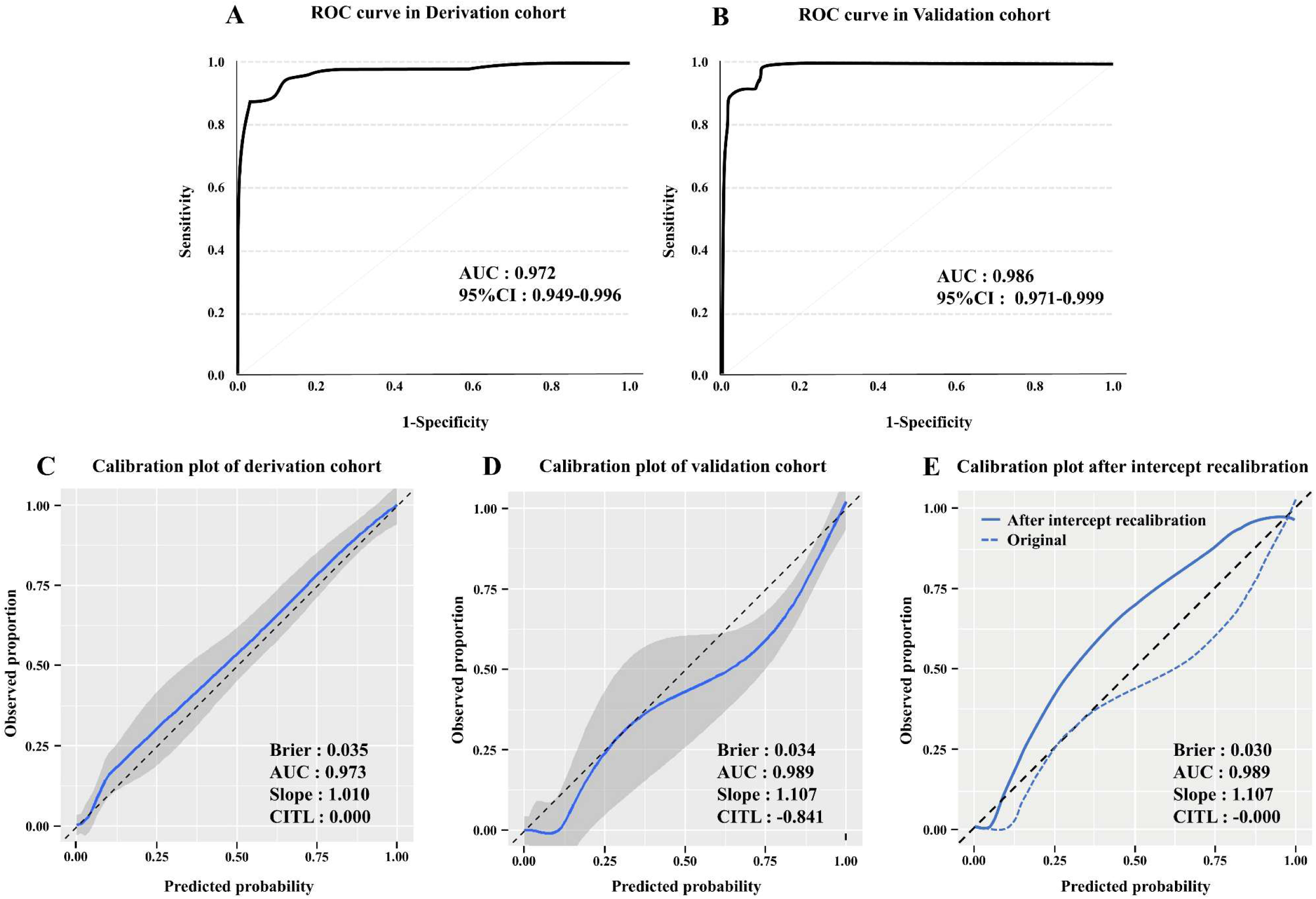
Performance of the prediction score and Calibration plots. Performance of the prediction score for the diagnosis of immunoglobulin light chain cardiac amyloidosis versus transthyretin cardiac amyloidosis in the derivation cohort (A) and validation cohort (B). Calibration plots between predicted and observed AL-CA in the derivation (C), validation (D) cohorts. Calibration plot after intercept-only recalibration in the validation cohort (E). Gray shading represents the 95% confidence band around the smoothed calibration curve. The dashed diagonal line represents the line of perfect calibration, and the solid blue line represents the smoothed calibration curve. ROC, receiver operating characteristic; AUC, area under the curve; CI, confidence interval.

### Calibration and Decision Curve Analysis

Calibration plots in the derivation cohort showed good agreement between predicted and observed probabilities (CITL 0.00; slope 1.01). In the validation cohort, discrimination remained preserved, but CITL was negative (CITL −0.84; slope 1.11), indicating systematic overestimation of predicted probability. After recalibration by updating the intercept, calibration improved (slope 1.09; CITL 0.00), with a Brier score of 0.03 and reduced overestimation in the intermediate to high probability range (Figure 2C, 2D, 2E).

In the derivation cohort, DCA demonstrated that the model provided consistently higher net benefit than treat-all and treat-none strategies across the entire range of threshold probabilities from 0.01 to 0.50 (Figure 3A). Similar findings were observed in the validation cohort. Net benefit was preserved after recalibration, indicating that improved calibration did not compromise clinical utility (Figure 3B).

**Figure 3.**
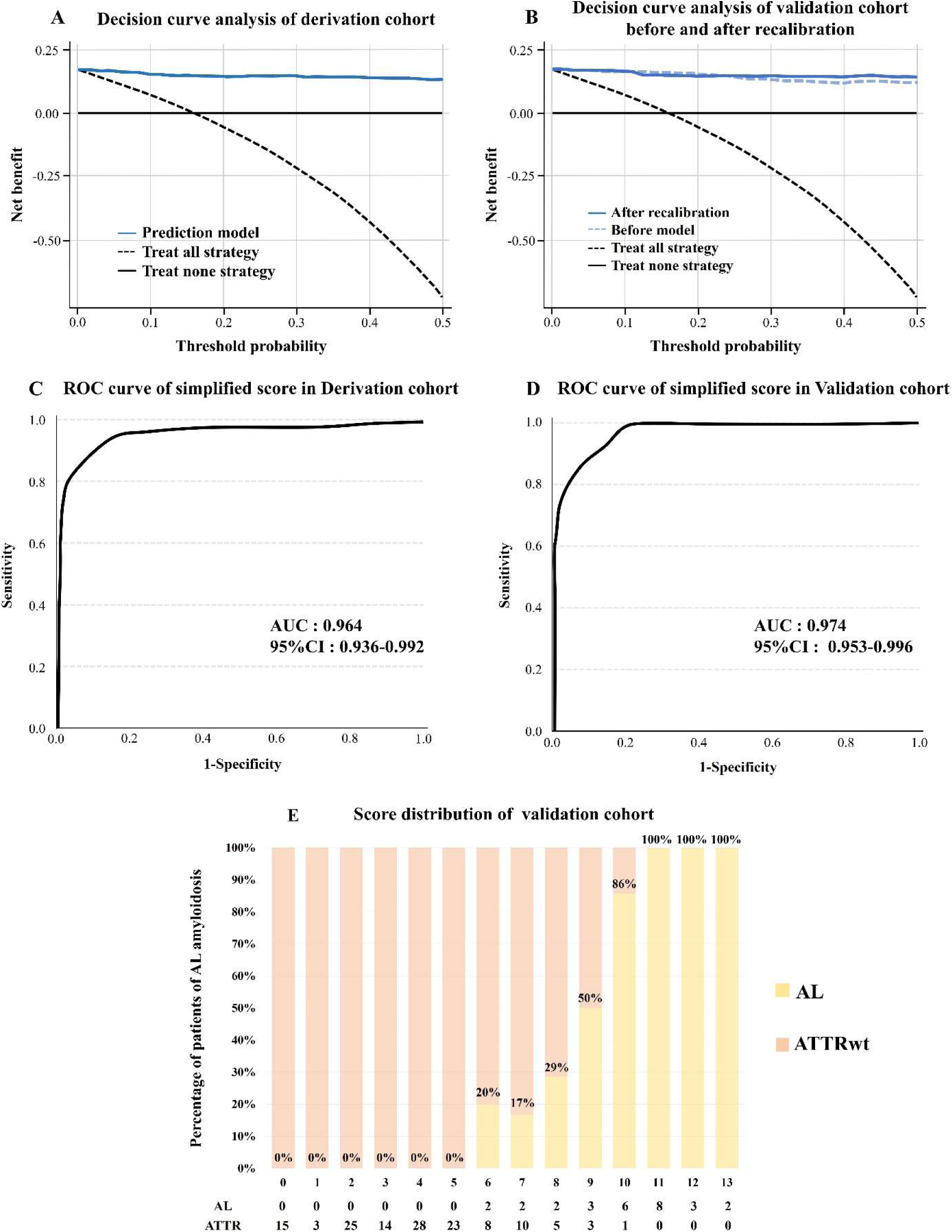
Decision curve analysis and performance of simplified score. Decision curve analysis showing clinical net benefit across threshold probabilities of the derivation cohort (A), and validation cohort before and after recalibration (B). Performance of the simplified score for the diagnosis of AL-CA in the derivation cohort (C) and validation cohort (D). The percentage of AL-CA (yellow bars) across the distribution of simplified score (E). AL, light-chain amyloidosis; ATTRwt, wild-type transthyretin amyloidosis; CA, cardiac amyloidosis; ROC, receiver operating characteristic; AUC, area under the curve; CI, confidence interval.

### Simplified Score Performance

A simplified scoring system was constructed by assigning points to the seven parameters (Table 2): age <74 years (2 points), absence of AF (2 points), absence of CTS (3 points), albumin <3.6 g/dL (1 point), proteinuria (2 points), low voltage (1 point), and LVPWT <14.2 mm (2 points). Total scores ranged from 0 to 13. The simplified score yielded an AUC of 0.96 (95% CI: 0.94–0.99) in the derivation cohort and 0.97 (95% CI: 0.95–0.99) in the validation cohort (Figure 3C, 3D).

Diagnostic accuracy at various cutoff points is shown in Table 3. The distribution of the simplified score and the corresponding proportion of AL-CA cases is shown in Figure 3E. Patients were categorized into three probability groups: low, intermediate, and high (Figure 4). In the validation cohort, 66% of patients were classified into the low- probability (0% confirmed AL-CA), while 95% of those in the high-probability group were confirmed to have AL-CA.

**Figure 4.**
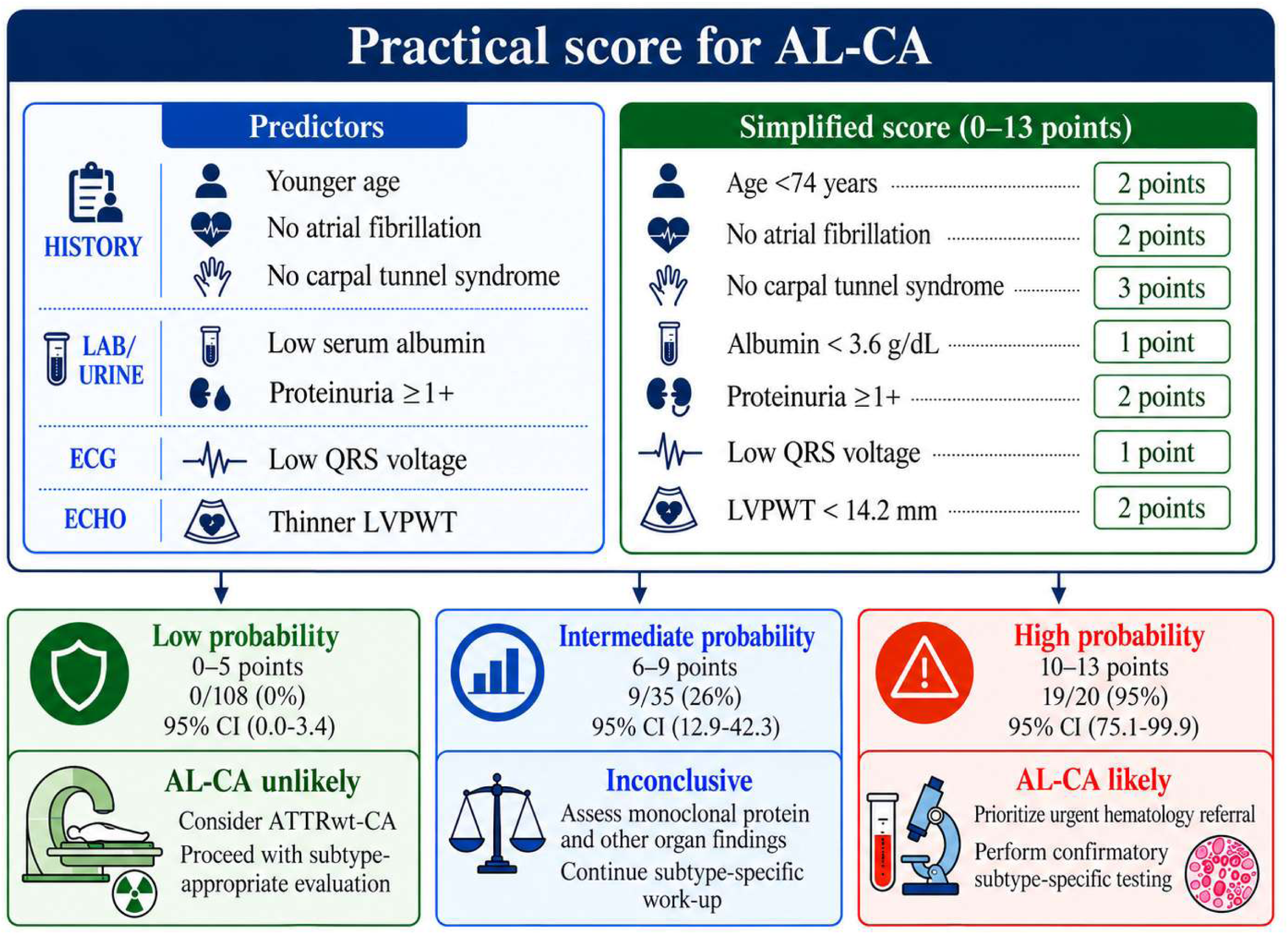
Clinical application of the simplified score for triage of suspected AL-CA. The score is intended to complement, not replace, standard monoclonal protein testing and definitive subtype-specific evaluation. In the validation cohort, no AL-CA cases were observed among patients scoring 0–5 points, whereas 19 of 20 patients scoring 10–13 points had AL-CA. These risk strata represent internal-validation estimates and require external validation before clinical implementation. AL-CA indicates immunoglobulin light-chain cardiac amyloidosis; ATTRwt-CA, wild-type transthyretin cardiac amyloidosis; CI, confidence interval; and LVPWT, left ventricular posterior wall thickness.

**Table 3.**
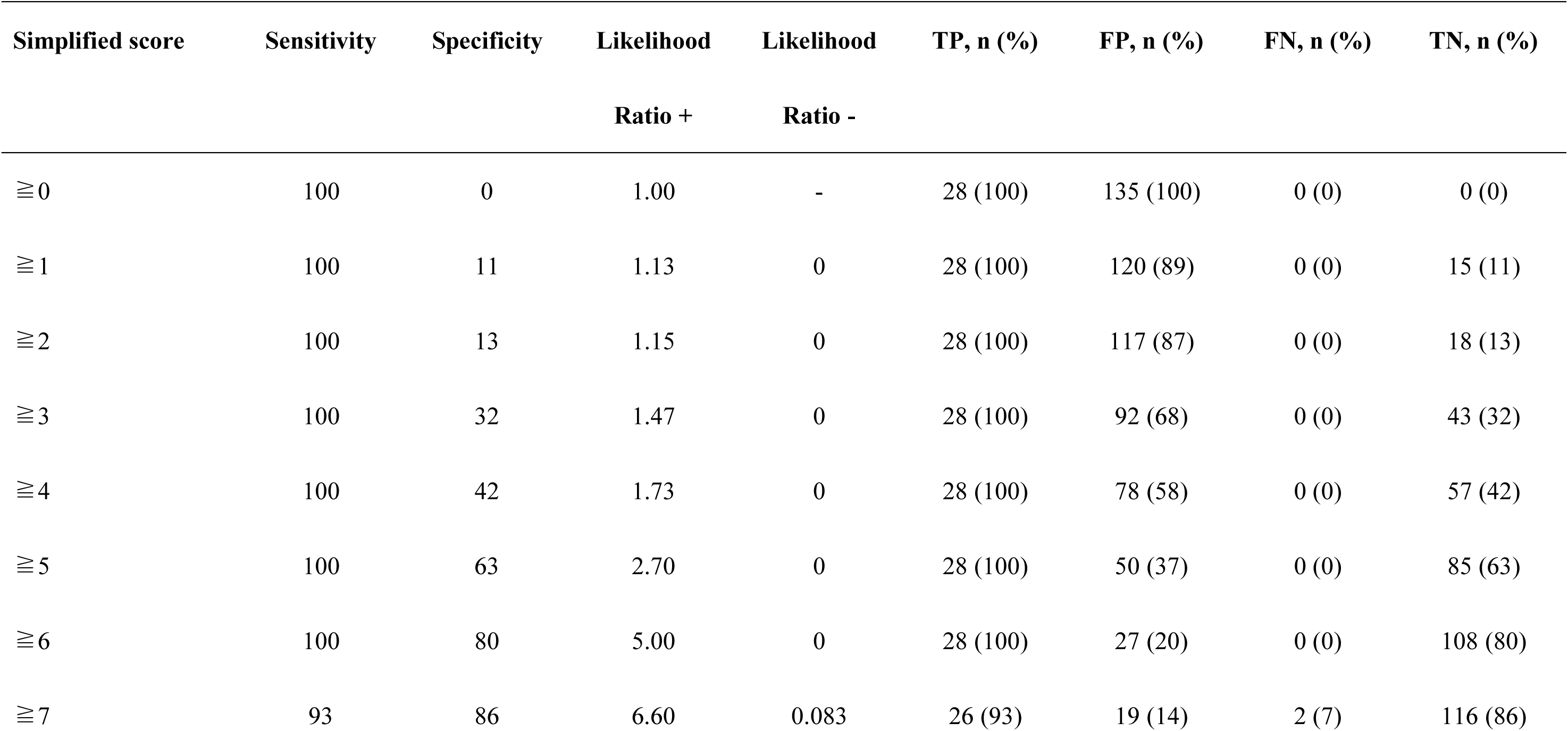
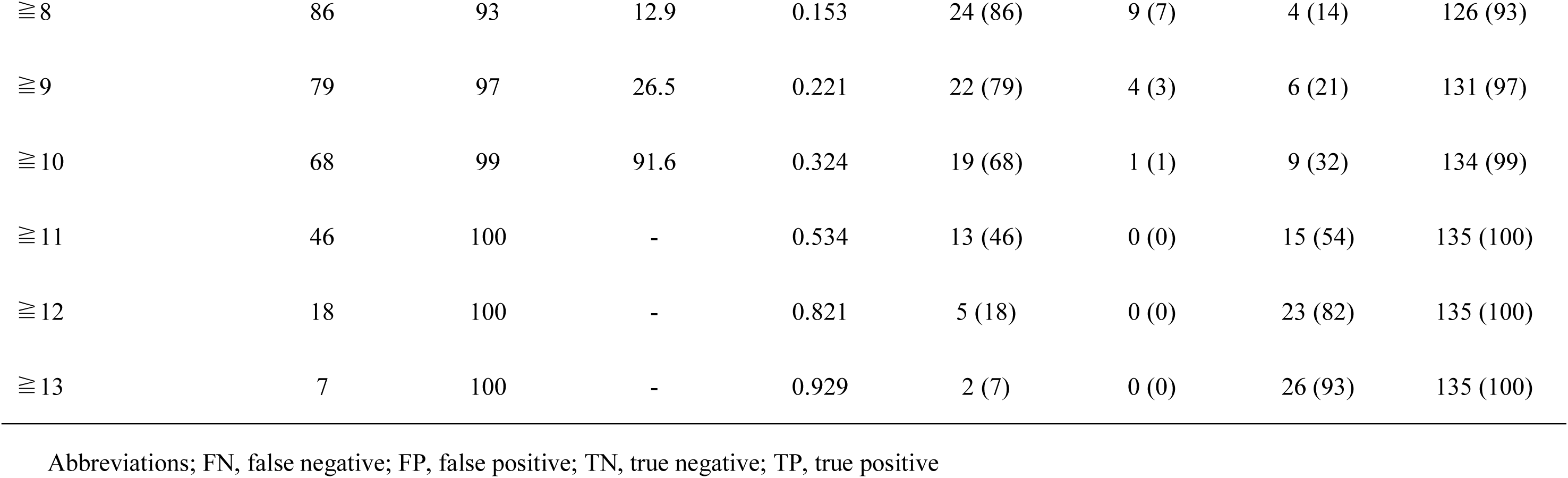
Sensitivity and specificity of the simplified score in identifying patients with AL-CA at different cutoffs.

To evaluate the incremental value of the simplified score, discriminative performance was compared across models: the AUC was 0.79 (95% CI, 0.69–0.87) for the FLC model, 0.82 (95% CI, 0.73–0.91) for the immunofixation model, 0.92 (95% CI, 0.85–0.98) for the score model (≥8 points), and 0.97 (95% CI, 0.92–1.00) for the combined model. The combined model significantly outperformed both the FLC model (ΔAUC, 0.19; 95% CI, 0.10–0.27; p<0.001) and the immunofixation model (ΔAUC, 0.14; 95% CI, 0.056–0.23; p = 0.001). Furthermore, compared with the dichotomized score model, the combined model improved reclassification metrics (continuous NRI, 1.29; p<0.001; IDI, 0.081; p=0.023), whereas the increase in AUC did not reach statistical significance (ΔAUC, 0.047; 95% CI, −0.002 to 0.096; p=0.062) (Table 4). In a sensitivity analysis restricted to patients with AL-CA and those with monoclonal protein–positive ATTRwt-CA, the simplified score (≥8 points) maintained excellent discrimination, with an AUC of 0.90 (95% CI, 0.82–0.99). At a cutoff of ≥8 points, the score identified AL-CA with a sensitivity of 86% and a specificity of 89% (Supplemental Table 4). The AUC of the simplified score at a cutoff of ≥8 points was significantly higher than that of the FLC model (ΔAUC, 0.37; 95% CI, 0.22–0.51; p<0.001) and the immunofixation model (ΔAUC, 0.21; 95% CI, 0.055–0.37; p = 0.008). Continuous NRI and IDI analyses also favored the score over both comparator tests (Supplemental Table 5).

**Table 4.** Discriminative performance and incremental predictive value of diagnostic models for AL amyloidosis.

**Panel A. Discriminative performance of each model**
| Model | Variables included | AUC (95% CI) |
| --- | --- | --- |
| FLC model | FLC abnormality | 0.79 (0.69–0.87) |
| Immunofixation model | Immunofixation positivity | 0.82 (0.73–0.91) |
| Score model | Simplified score $\geq 8$ | 0.92 (0.85–0.98) |
| Combined model | FLC abnormality, immunofixation positivity, and diagnostic score | 0.97 (0.92–1.00) |

**Panel B. Pairwise comparison of model performance**
| Reference model | New model | ΔAUC (95% CI) | DeLong<br>p-value | Continuous<br>NRI | p-value | IDI | p-value |
| --- | --- | --- | --- | --- | --- | --- | --- |
| FLC model | Immunofixation model | 0.044 (-0.052–0.14) | 0.37 | 0.099 | 0.49 | 0.17 | 0.001 |
| FLC model | Score model | 0.14 (0.032–0.25) | 0.011 | 1.67 | <0.001 | 0.44 | <0.001 |
| FLC model | Combined model | 0.19 (0.10–0.27) | <0.001 | 1.59 | <0.001 | 0.52 | <0.001 |
| Immunofixation model | Score model | 0.096 (-0.014–0.21) | 0.087 | 1.67 | <0.001 | 0.27 | <0.001 |
| Immunofixation model | Combined model | 0.14 (0.056–0.23) | 0.001 | 1.67 | <0.001 | 0.35 | <0.001 |
| Score model | Combined model | 0.047 (-0.002–0.096) | 0.062 | 1.29 | <0.001 | 0.081 | 0.023 |

Abbreviations; AL, immunoglobulin light-chain amyloidosis; AUC, area under the receiver operating characteristic curve; CI, confidence interval; FLC, free light-chain; IDI, integrated discrimination improvement; NRI, net reclassification improvement.
Notes: Analyses were performed in the common complete-case population of 129 patients, including 26 patients with AL-CA. AUC confidence intervals and pairwise differences in AUC were estimated using DeLong's method. ΔAUC was calculated as the AUC of the new model minus that of the reference model. NRI was calculated as continuous, category-free NRI. NRI and IDI p values were based on asymptotic Wald tests

## Discussion

### Principal Findings and Clinical Positioning of the Score

In this study, we developed and internally validated a simple seven-variable score composed of age, history of AF, history of CTS, serum albumin, proteinuria, low voltage on electrocardiography, and LVPWT to detect AL-CA among patients undergoing evaluation for suspected cardiac amyloidosis. Although this was a single- center study, the model was tested in an internal validation cohort from a large consecutive real-world population. The model showed excellent discrimination in both cohorts, and the simplified score preserved similarly high performance. The high AUC values observed in both cohorts must be interpreted appropriately. Because AL-CA and ATTRwt-CA are pathophysiologically distinct diseases, the high discrimination may reflect the spectrum bias between the two subtypes when appropriate variables are measured. The clinical value of this score lies not only in the AUC but also in the fact that a high level of discrimination can be achieved without the need for advanced imaging tests or biopsy. In addition, Decision curve analysis demonstrated positive net benefit across a wide range of threshold probabilities compared with treat-all and treat- none strategies. Given the substantial clinical consequences of delayed AL-CA diagnosis, the observed benefit at relatively low threshold probabilities supports the potential use of the model as an early triage tool. However, these thresholds should not be interpreted as specific cutoffs for individual diagnostic procedures.

### Clinical Utility in Real-World Practice

Because diagnostic delay in AL-CA directly affects disease stage and survival (9), a first-visit triage tool based on readily available variables may enable cardiologists to initiate subtype-oriented evaluation before advanced imaging or invasive testing is completed.

Previously reported approaches have focused either on detecting cardiac amyloidosis itself rather than distinguishing AL-CA from ATTRwt-CA (23), or on subtype differentiation using advanced echocardiographic or cardiac magnetic resonance parameters (24). By contrast, this score was designed to use variables with low acquisition burden and broad real-world availability. This may facilitate implementation across institutions with varying resources, particularly when time-sensitive triage is needed.

Importantly, this score is not intended to replace multimodality imaging or specialized diagnostic testing. Imaging-based approaches are central to the diagnosis of CA (3, 25), but they may not always be immediately available in routine practice and are not immune to false-positive findings (26). Rather, this score may serve as an early clinical gatekeeper: a tool applied at the first encounter to identify which patients should undergo expedited hematologic assessment, advanced imaging, and definitive diagnostic workup.

The most useful feature of the score may be its ability to support risk-based triage. Within a population already undergoing evaluation for CA, a low score effectively excluded AL-CA in the validation cohort, whereas intermediate or high scores should prompt its active exclusion, and high scores warrant urgent hematology collaboration (Figure 4). An additional notable feature of the present score is its diagnostic performance relative to conventional monoclonal protein testing for AL-CA. Although an abnormal κ/λ ratio and positive urine and serum immunofixation electrophoresis represent standard screening tests for AL-CA, discrimination using the simplified score was maintained even in the difficult-to-distinguish cohort comprising patients with AL-CA and those with ATTRwt-CA who tested positive for these assays (Supplemental Table 4). Despite advances in multimodality imaging, the coexistence of monoclonal gammopathy remains an important diagnostic challenge in patients with suspected ATTRwt-CA. Our findings suggest that the addition of this score to these monoclonal protein tests provides incremental value in terms of improving the diagnostic accuracy of AL-CA. This score may help prioritize patients who require urgent hematologic evaluation and tissue-based amyloid typing.

### Biological Plausibility of the Selected Predictors

The seven variables are not only statistically robust but also reflect distinct pathophysiological mechanisms that differentiate AL-CA from ATTRwt-CA. In our cohort, older age, history of AF, and history of CTS were each negatively associated with AL-CA, consistent with their well-established roles as hallmark “red flags” of ATTRwt-CA (27). The rarity of CTS and AF in our patients with AL-CA, together with their high frequency in those with ATTRwt-CA, underscores the diagnostic value of careful history taking at first presentation.

Our data also suggest that AL-CA presents with a combination of greater systemic involvement and less advanced structural thickening at first evaluation. Proteinuria and hypoalbuminemia reflect the systemic nature of AL amyloidosis. Renal involvement is observed in two-thirds of patients with AL amyloidosis, and the causes include amyloid deposition in the glomeruli and direct nephrotoxicity caused by light chains (28).

Notably, in our cohort, proteinuria differed between subtypes, but the difference in eGFR was small, and it was not selected in the multivariate analysis. This pattern suggests that, at least at the time of first cardiology assessment, renal involvement in AL amyloidosis may be captured more sensitively by urinary protein loss than by overt reduction in filtration function. In contrast, eGFR may be influenced by age-related or heart failure-related renal dysfunction in older ATTRwt-CA patients, thereby reducing its discriminatory value in this setting. The coexistence of relatively thinner LVPWT and frequent low voltage suggests that AL-CA may cause substantial myocardial dysfunction and electrical impairment before marked wall thickening develops. This interpretation is also compatible with the concept that circulating light chains exert direct cardiotoxic effects in addition to tissue infiltration (29–31).

These findings suggest that the systemic footprint of AL-CA is detectable through routine laboratory parameters well before advanced cardiac involvement is apparent. This key point is often underutilized in routine cardiology practice.

### Statistical Approaches and Clinical Advantage

These statistical approaches were chosen to ensure reliability for clinical use rather than to maximize apparent performance. The consistent selection of the same seven variables across all imputed datasets supports the robustness of the predictors, and the miscalibration observed in the validation cohort was confined to baseline prevalence and correctable by intercept updating alone. DCA further indicated that the score minimizes the most consequential error—missing AL-CA—at low threshold probabilities.

### Study Limitations

This study has several limitations. First, it was a retrospective, single-center study and is therefore subject to selection bias, measurement bias, and center-specific referral patterns. Second, the long enrollment period also spanned substantial changes in referral patterns, monoclonal protein testing, multimodality imaging, and diagnostic algorithms, which may have introduced temporal heterogeneity. Third, calibration may drift over time or across institutions because of changes in prevalence and practice patterns; although the degree of miscalibration in the present study was modest and was corrected by intercept updating alone, external validation is essential before widespread implementation. Fourth, the score was not evaluated in broader populations such as patients with unexplained left ventricular hypertrophy or HFpEF and its performance in these clinically important groups requires further study. Finally, although DCA suggests potential clinical usefulness, prospective impact studies are required to determine whether implementation of this score shortens diagnostic delay and improves survival, treatment timing, or quality of life.

## Conclusions

We developed a practical score for differentiating AL-CA from ATTRwt-CA using only variables at the first visit. This score has the potential to serve as a practical early triage tool for identifying patients who require rapid evaluation for AL-CA. External validation and prospective implementation studies are needed to determine whether adopting this approach will reduce diagnostic delays and improve clinically meaningful outcomes. This score is not a standalone diagnostic test and is best used alongside monoclonal protein testing.

## Data Availability

The data are not publicly available because they contain information that could compromise participant privacy; deidentified data may be available from the corresponding author upon reasonable request and institutional approval.

## Acknowledgements

The authors thank all cardiologists in Kumamoto University Hospital who contributed to the clinical care of the patients included in this study and to data collection for the analysis.

## Abbreviations

CA: cardiac amyloidosis
AL-CA: immunoglobulin light-chain cardiac amyloidosis
ATTRwt-CA: wild-type transthyretin cardiac amyloidosis
HFpEF: heart failure with preserved ejection fraction
FLC: free light chain
AF: atrial fibrillation
CTS: carpal tunnel syndrome
TRIPOD: Transparent Reporting of a multivariable prediction model for Individual Prognosis Or Diagnosis
MICE: multiple imputation by chained equations
LASSO: least absolute shrinkage and selection operator
AUC: area under the receiver operating characteristic curve
CITL: calibration-in-the-large
DCA: decision curve analysis
NRI: net reclassification improvement
IDI: integrated discrimination improvement
LVPWT: left ventricular posterior wall thickness
CI: confidence interval.

---

**Supplemental Table 1.**
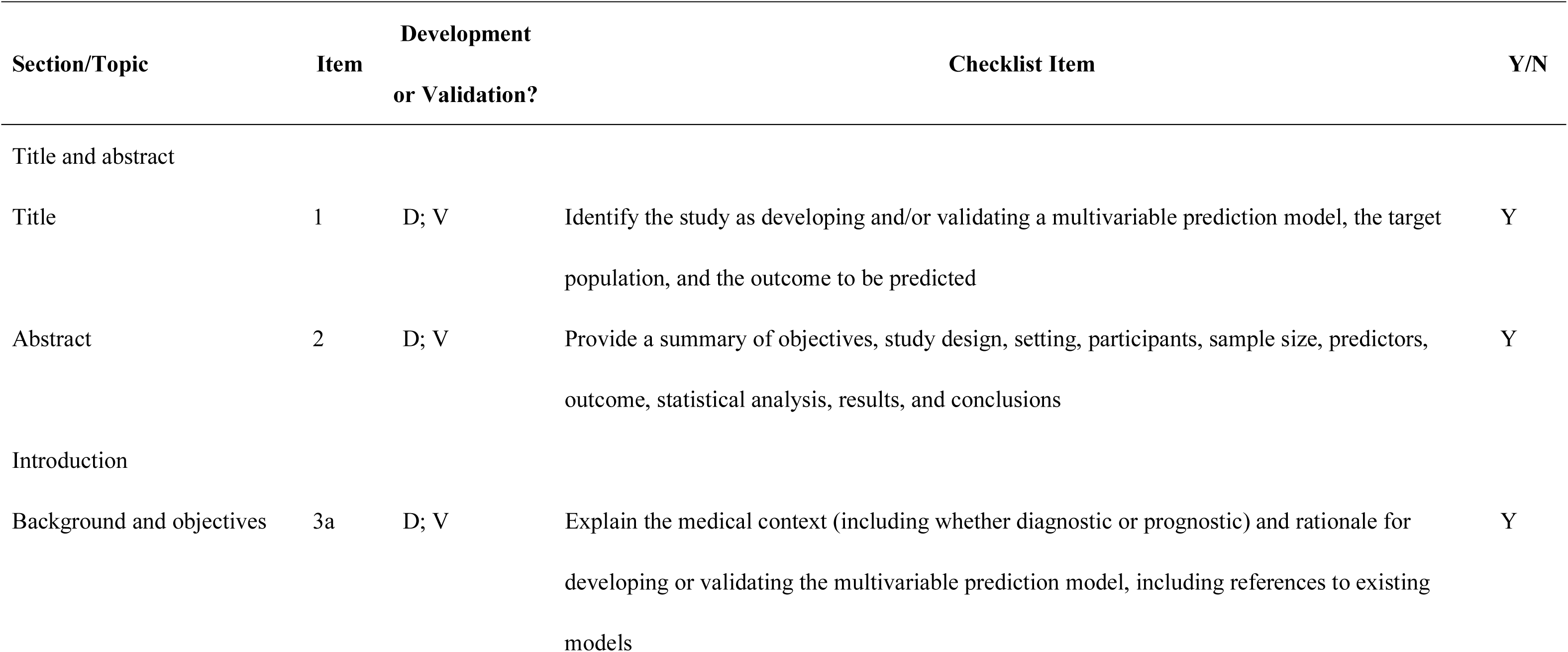
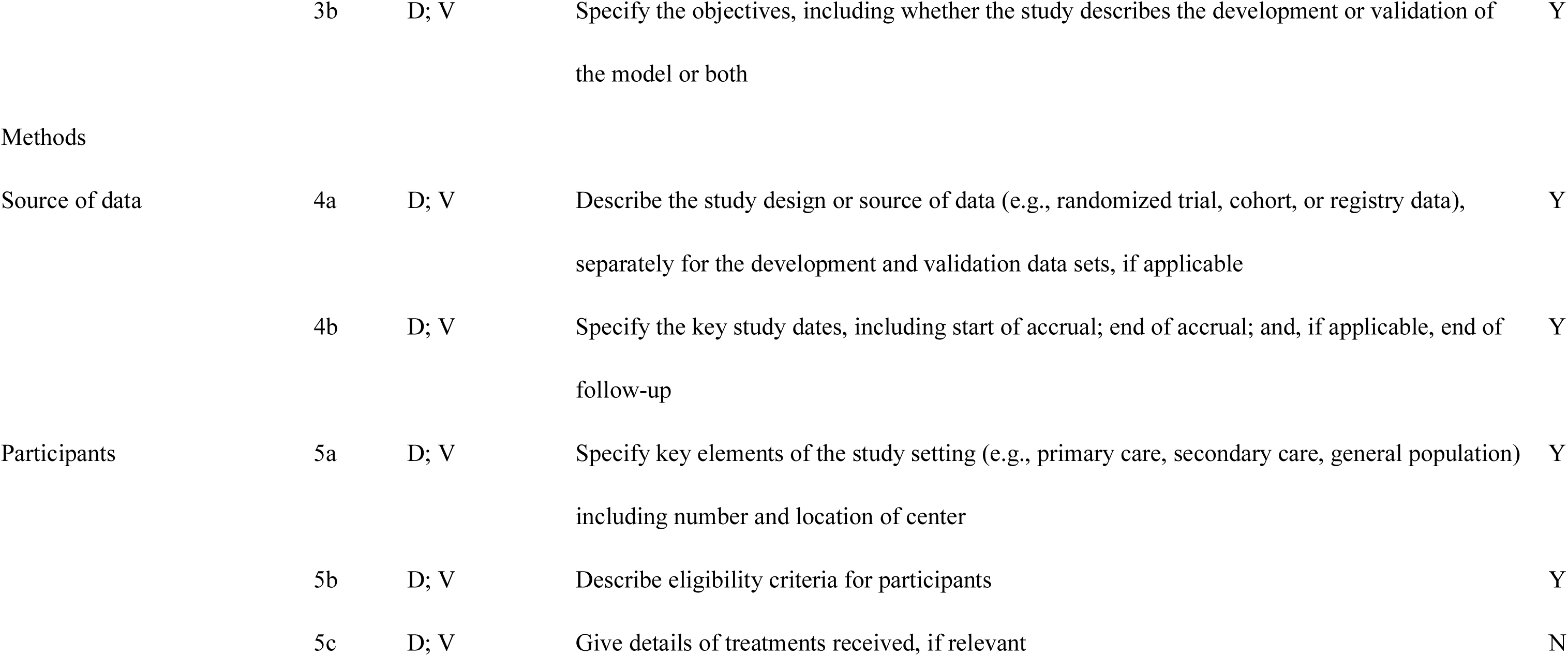
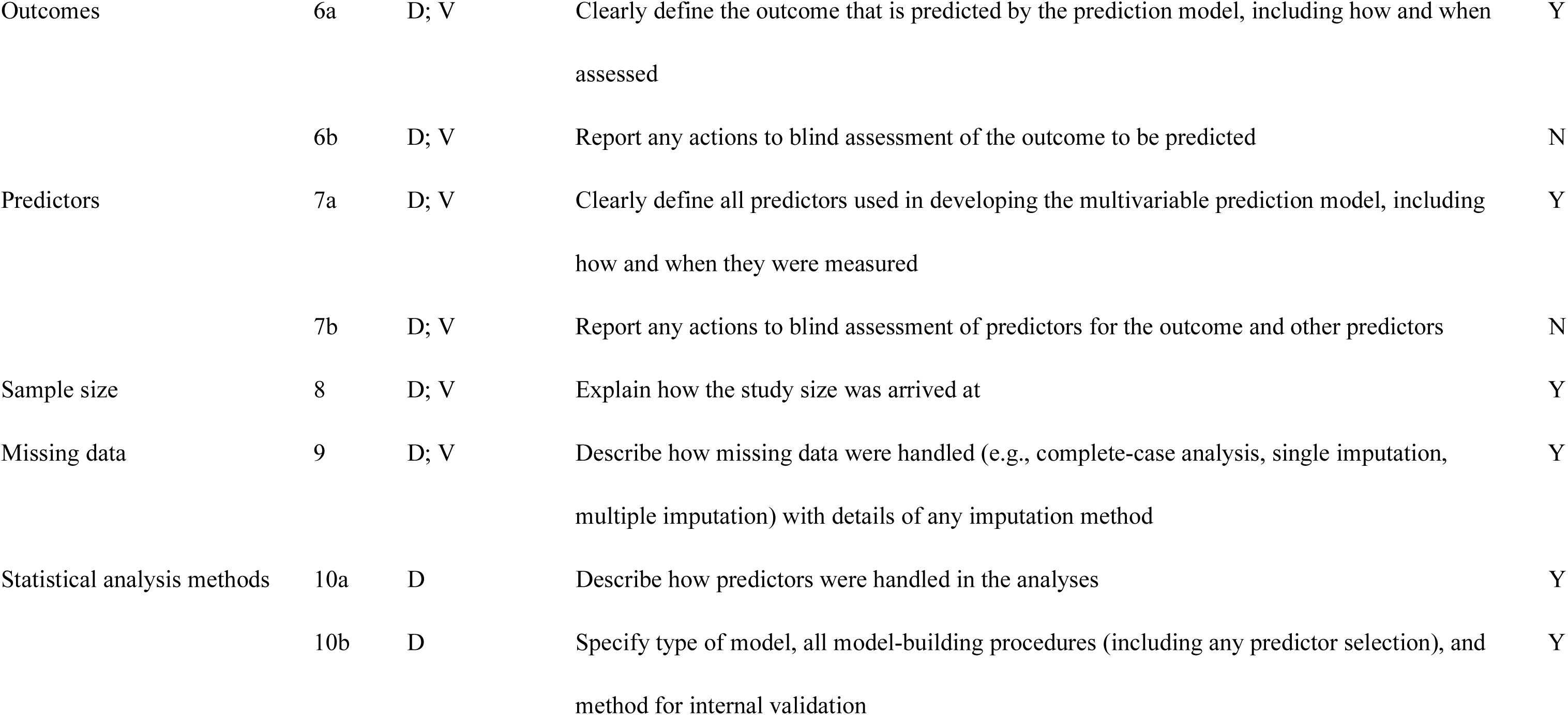
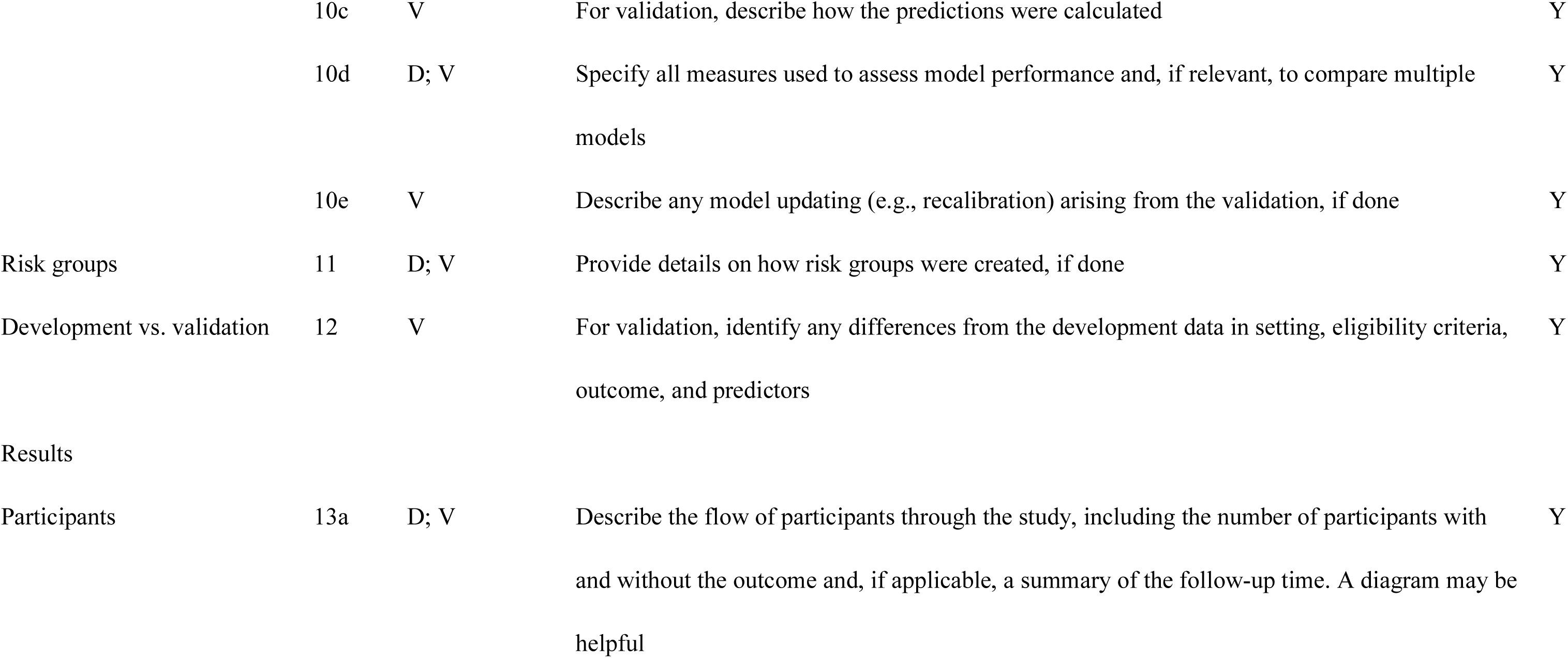
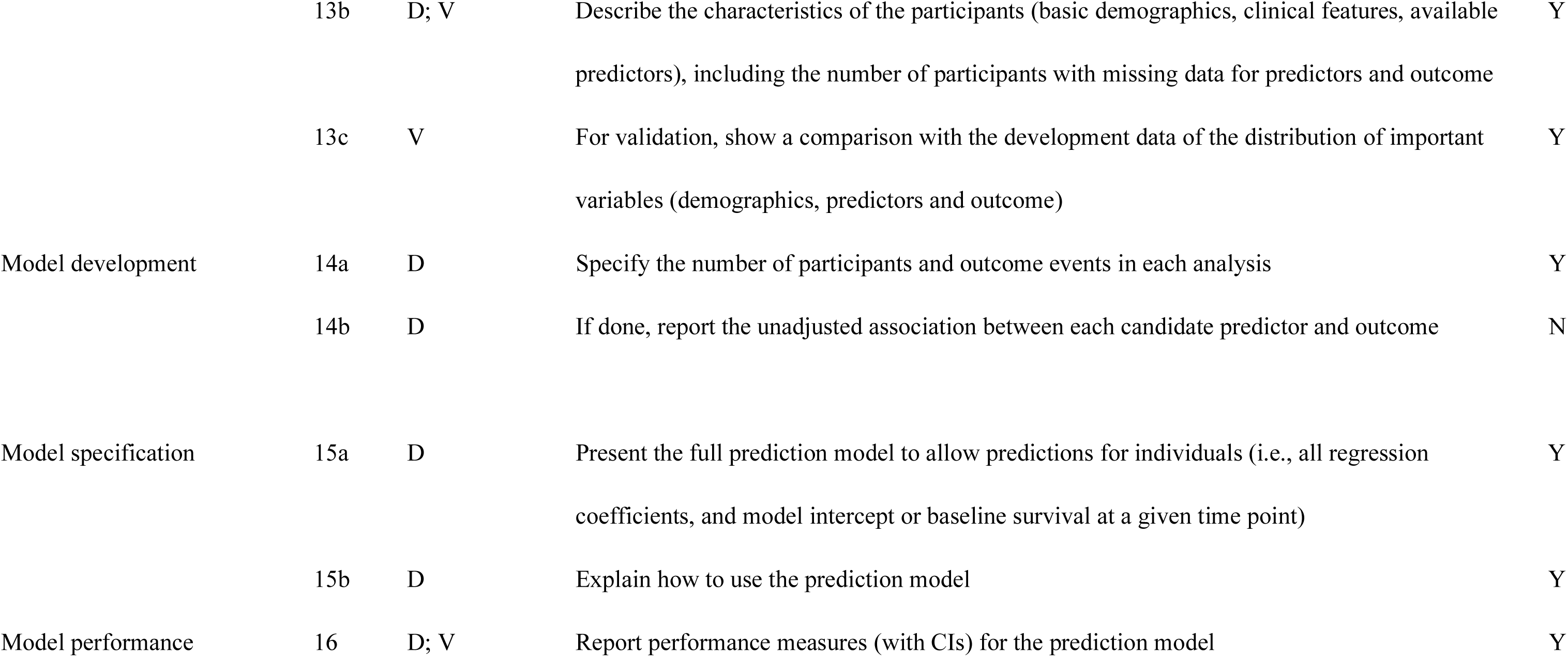
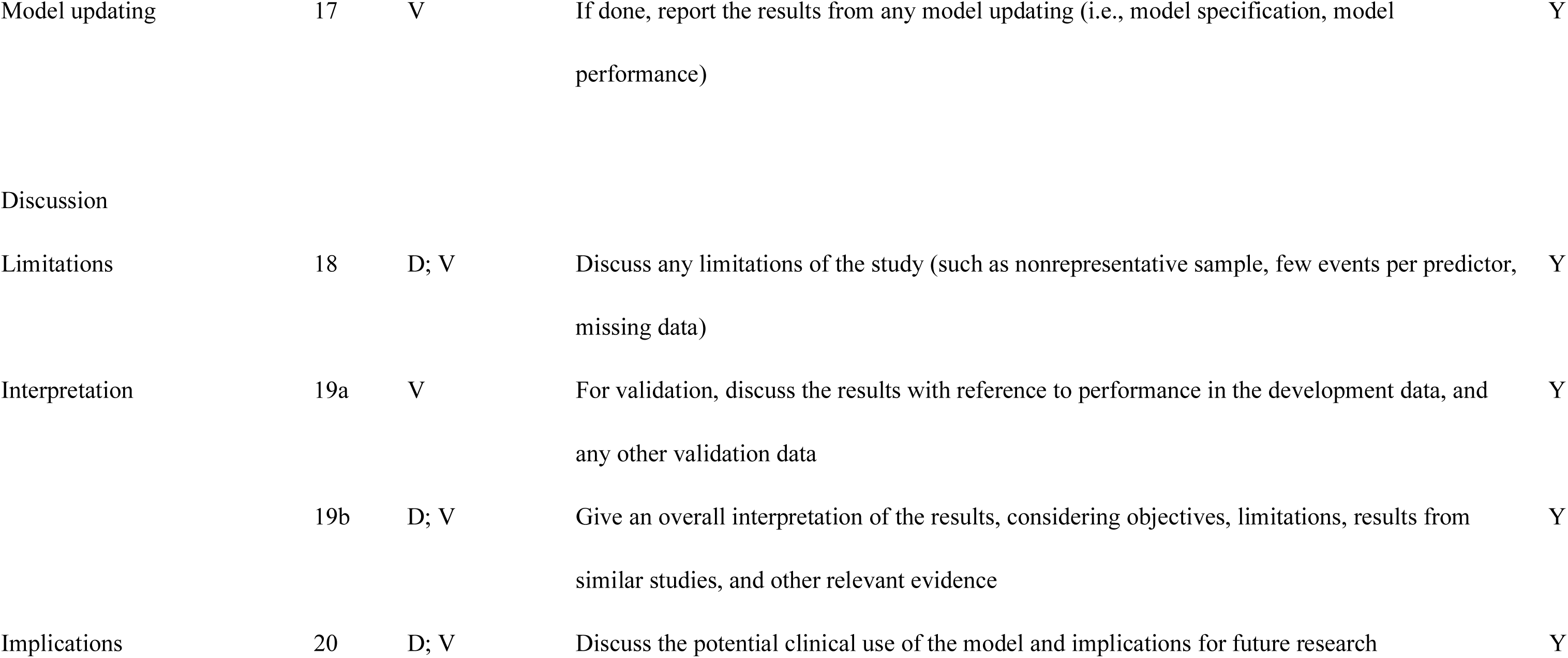
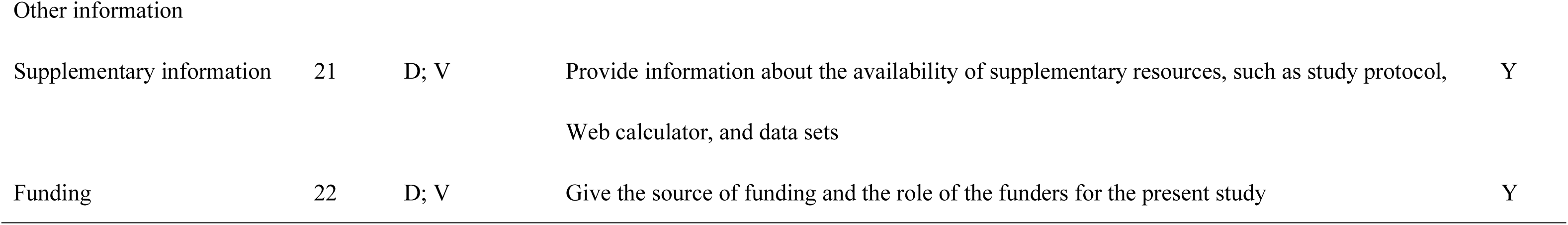
Checklist of TRIPOD items.

**Supplemental Table 2.** Patient baseline characteristics of the validation cohort.

|  | Overall | AL | ATTRwt | missing | p-value |
| --- | --- | --- | --- | --- | --- |
| Total | 163 | 28 | 135 | 0 |  |
| **Age (years)** | **75.3 ± 8.66** | **66.0 ± 10.4** | **77.3 ± 6.84** | **0** | **<0.001** |
| **Male** | **131 (80)** | **15 (54)** | **116 (86)** | **0** | **<0.001** |
| **Body mass index (kg/m<sup>2</sup>)** | 22.7 ± 3.00 | 22.2 ± 3.22 | 22.8 ± 2.96 | 0 | 0.326 |
| Comorbidity |  |  |  |  |  |
| **Hypertension** | **94 (58)** | **11 (40)** | **83 (61)** | **0** | **0.030** |
| Diabetes mellitus | 43 (26) | 7 (25) | 36 (27) | 0 | 0.855 |
| Dyslipidemia | **57 (35)** | **5 (18)** | **52 (39)** | **0** | **0.037** |
| Chronic kidney disease | 99 (61) | 15 (54) | 84 (62) | 0 | 0.394 |
| Hemodialysis | 3 (2) | 1 (4) | 2 (2) | 0 | 0.434 |
| **Atrial fibrillation** | **79 (49)** | **4 (14)** | **75 (56)** | **0** | **<0.001** |
| Ischemic heart disease | 17 (10) | 3 (11) | 14 (10) | 0 | 1.0 |
| **Carpal tunnel syndrome** | **71 (44)** | **1 (4)** | **70 (52)** | **0** | **<0.001** |
| **Spinal canal stenosis** | **42 (26)** | **0 (0)** | **42 (31)** | **0** | **0.002** |
| Permanent pacemaker | 16 (10) | 4 (14) | 12 (9) | 0 | 0.482 |
| Implantable cardioverter defibrillator | 4 (3) | 2 (7) | 2 (2) | 0 | 0.137 |
| **Post radiofrequency catheter ablation** | **25 (15)** | **1 (4)** | **25 (18)** | **0** | **0.081** |
| Post PCI / CABG | 18 (11) | 2 (7) | 16 (12) | 0 | 0.54 |
| History of heart failure | 80 (49) | 13 (46) | 67 (50) | 0 | 0.837 |
| **Hospitalization of heart failure** | **52 (32)** | **10 (36)** | **42 (31)** | **0** | **0.66** |
| **NYHA functional class** |  |  |  | **0** | **0.278** |
| I | 33 (20) | 7 (25) | 26 (19) |  |  |
| II | 78 (48) | 9 (32) | 69 (51) |  |  |
| III | 49 (30) | 11 (39) | 38 (28) |  |  |
| IV | 3 (2) | 1 (4) | 2 (2) |  |  |
| **NYHA II or more** | **130 (80)** | **21 (75)** | **109 (81)** | **0** | **0.605** |
| **Monoclonal gammopathy** | **55 (34)** | **28 (100)** | **27 (20)** | **0** | **<0.001** |
| **Serum free $\kappa$ light chain (mg/L)** | **27.6 (17.4–50.1)** | **20.6 (12.3–66.3)** | **27.8 (20.4–46.3)** | **33** | **<0.001** |
| **Serum free $\lambda$ light chain (mg/L)** | **23.7 (17.8–44.1)** | **190.0 (49.8–558.5)** | **22.6 (17.2–31.8)** | **33** | **<0.001** |
| **Abnormal $\kappa$ / $\lambda$ ratio** | **42 (26)** | **20 (71)** | **22 (16)** | **33** | **<0.001** |
| **Positive serum and / or urine immunofixation** | **30 (18)** | **20 (71)** | **10 (7)** | **22** | **<0.001** |
| Electrocardiographic parameters |  |  |  |  |  |
| **PR duration (ms)** | 200.1 ± 51.8 | 188.0 ± 41.7 | 203.8 ± 54.2 | 56 | 0.184 |
| **QRS duration (ms)** | **111.3 ± 27.8** | **95.3 ± 15.3** | **114.9 ± 28.7** | **13** | **<0.001** |
| LBBB | 16 (11) | 1 (4) | 15 (12) | 13 | 0.308 |
| RBBB | **26 (17)** | **1 (4)** | **25 (21)** | **13** | **0.049** |
| **QTc (ms)** | **455.5 ± 33.8** | **438.6 ± 36.9** | **459.3 ± 32.0** | **13** | **0.003** |
| **Low QRS voltage** | **28 (18)** | **17 (61)** | **11 (8)** | **13** | **<0.001** |
| Pseudo-infarct pattern | 89 (56) | 17 (61) | 72 (55) | 13 | 0.578 |
| Echocardiographic parameters |  |  |  |  |  |
| **IVST (mm)** | **14.9 ± 2.68** | **13.5 ± 2.75** | **15.2 ± 2.57** | **0** | **0.002** |
| **LVPWT (mm)** | **15.1 ± 2.89** | **13.5 ± 2.76** | **15.4 ± 2.82** | **0** | **0.001** |
| **LVEF (%)** | **52.8 ± 9.86** | **59.0 ± 8.66** | **51.5 ± 9.62** | **0** | **<0.001** |
| LVEDD (mm) | 41.6 ± 6.39 | 39.3 ± 6.10 | 42.0 ± 6.36 | 0 | 0.035 |
| **LVESD (mm)** | **31.1 ± 6.75** | **27.0 ± 6.89** | **31.9 ± 6.43** | **0** | **0.001** |
| **LVMI (g/m<sup>2</sup>)** | **156.8 ± 42.1** | **125.9 ± 36.1** | **163.2 ± 40.4** | **0** | **<0.001** |
| **Left atrial diameter (mm)** | **41.7 ± 7.83** | **37.9 ± 8.44** | **42.5 ± 7.48** | **0** | **0.004** |
| Mitral deceleration time (ms) | 183.0 ± 59.3 | 190.8 ± 74.2 | 181.4 ± 56.0 | 0 | 0.443 |
| E/A | 1.80 ± 1.34 | 1.55 ± 1.03 | 1.88 ± 1.43 | 41 | 0.205 |
| **E/e'** | 20.0 ± 7.20 | 21.7 ± 8.07 | 19.7 ± 6.99 | 0 | 0.177 |
| Pericardial effusion | 77 (48) | 14 (52) | 63 (47) | 0 | 0.646 |
| Pleural effusion | **39 (24)** | **13 (48)** | **26 (19)** | **0** | **0.001** |
| Moderate mitral regurgitation or more | 18 (11) | 2 (7) | 16 (12) | 0 | 0.741 |
| Moderate aortic stenosis or more | 12 (7) | 0 (0) | 12 (9) | 0 | 0.129 |
| Laboratory values |  |  |  |  |  |
| BNP (pg/mL) | **180.9 (100.2–405.6)** | **305.9 (156.6–714.0)** | **160.9 (97.7–330.4)** | **2** | **0.008** |
| hs-troponin T (ng/mL) | 0.052 (0.035–0.086) | 0.057 (0.034–0.171) | 0.050 (0.035–0.078) | 2 | 0.365 |
| Creatinine (mg/dL) | 1.21 ± 0.79 | 1.41 ± 1.24 | 1.17 ± 0.66 | 0 | 0.332 |
| eGFR (mL/min/1.73 m<sup>2</sup>) | 52.9 ± 19.3 | 53.3 ± 26.4 | 52.8 ± 17.5 | 0 | 0.92 |
| **Hb (g/dL)** | 13.4 ± 1.93 | 12.9 ± 2.26 | 13.5 ± 1.85 | 0 | 0.136 |
| **Plt (×10<sup>3</sup>/μL)** | 181.6 ± 60.9 | 195.4 ± 86.9 | 178.7 ± 53.9 | 0 | 0.189 |
| **TP (g/dL)** | **6.84 ± 0.87** | **6.00 ± 1.25** | **7.02 ± 0.65** | **0** | **<0.001** |
| **Alb (g/dL)** | **3.86 ± 0.65** | **3.14 ± 1.00** | **4.02 ± 0.42** | **0** | **<0.001** |
| Na (mmol/L) | 139.6 ± 3.28 | 138.3 ± 4.01 | 139.8 ± 3.06 | 0 | 0.024 |
| K (mmol/L) | 4.37 ± 0.46 | 4.27 ± 0.42 | 4.40 ± 0.47 | 0 | 0.197 |
| Cl (mmol/L) | 104.3 ± 3.76 | 104.0 ± 4.56 | 104.3 ± 3.59 | 0 | 0.631 |
| PT (%) | 78.4 ± 29.9 | 86.6 ± 29.7 | 76.6 ± 29.8 | 8 | 0.11 |
| APTT (s) | 34.3 ± 9.97 | 32.6 ± 9.07 | 34.7 ± 10.2 | 8 | 0.315 |
| **Qualitative urine protein test** |  |  |  | **14** | **<0.001** |
| Negative | 110 (74) | 3 (12) | 107 (87) |  |  |
| Trace | 9 (6) | 2 (8) | 7 (6) |  |  |
| 1+ | 10 (7) | 5 (19) | 5 (4) |  |  |
| 2+ | 9 (6) | 7 (27) | 2 (2) |  |  |
| 3+ | 7 (5) | 5 (19) | 2 (2) |  |  |
| 4+ | 4 (3) | 4 (15) | 0 (0) |  |  |
| **Proteinuria** | **30 (20)** | **21 (81)** | **9 (7)** | **14** | **<0.001** |

Values are mean (SD), n / N (%), or median (IQR).
Abbreviations: AL = light-chain amyloidosis; ATTRwt = wild-type transthyretin amyloidosis; PCI = percutaneous coronary intervention; CABG = coronary artery bypass grafting; IVST = Interventricular septal thickness; LVPWT = Left ventricular posterior wall thickness; LVEF = Left ventricular ejection fraction; LVEDD = Left ventricular end-diastolic diameter; LVESD = Left ventricular end-systolic diameter; LVMI = Left ventricular mass index.

**Supplemental Table 3.** The result of Least Absolute Shrinkage and Selection Operator logistic regression.

| Variables | Selected datasets | Selection frequency | Missing rate |
| --- | --- | --- | --- |
| Age (years) | 20 / 20 | 100% | 0% |
| Alb (g/dL) | 20 / 20 | 100% | 0% |
| Atrial fibrillation | 20 / 20 | 100% | 0% |
| Carpal tunnel syndrome | 20 / 20 | 100% | 0% |
| Low QRS voltage | 20 / 20 | 100% | 2% |
| LVPWT (mm) | 20 / 20 | 100% | 0% |
| Proteinuria | 20 / 20 | 100% | 7% |
| Hb (g/dL) | 15 / 20 | 75% | 0% |
| LVEF (%) | 6 / 20 | 30% | 0% |
| Hemodialysis | 6 / 20 | 30% | 0% |
| Pseudo-infarct pattern | 1 / 20 | 5% | 2% |
| Male | 0 / 20 | 0% | 0% |
| Body mass index (kg/m<sup>2</sup>) | 0 / 20 | 0% | 0% |
| Hypertension | 0 / 20 | 0% | 0% |
| Diabetes mellitus | 0 / 20 | 0% | 0% |
| Dyslipidemia | 0 / 20 | 0% | 0% |
| Chronic kidney disease | 0 / 20 | 0% | 0% |
| Ischemic heart disease | 0 / 20 | 0% | 0% |
| Permanent pacemaker | 0 / 20 | 0% | 0% |
| Implantable cardioverter defibrillator | 0 / 20 | 0% | 0% |
| History of heart failure | 0 / 20 | 0% | 0% |
| Hospitalization of heart failure | 0 / 20 | 0% | 0% |
| NYHA II or more | 0 / 20 | 0% | 0% |
| QRS duration (ms) | 0 / 20 | 0% | 9% |
| LBBB | 0 / 20 | 0% | 9% |
| RBBB | 0 / 20 | 0% | 9% |
| QTc (ms) | 0 / 20 | 0% | 9% |
| Mitral deceleration time (ms) | 0 / 20 | 0% | 0% |
| E/e' | 0 / 20 | 0% | 0% |
| Pericardial effusion | 0 / 20 | 0% | 0% |
| Pleural effusion | 0 / 20 | 0% | 0% |
| Moderate mitral regurgitation or more | 0 / 20 | 0% | 0% |
| Moderate aortic stenosis or more | 0 / 20 | 0% | 0% |
| hs-troponin T (ng/mL) | 0 / 20 | 0% | 1% |
| Plt ($\times 10^3/\mu\text{L}$) | 0 / 20 | 0% | 0% |
| eGFR (mL/min/1.73 m<sup>2</sup>) | 0 / 20 | 0% | 0% |
| Na (mmol/L) | 0 / 20 | 0% | 0% |
| K (mmol/L) | 0 / 20 | 0% | 0% |
| Cl (mmol/L) | 0 / 20 | 0% | 0% |
| PT (%) | 0 / 20 | 0% | 8% |
| Spinal canal stenosis | VIF | - | 0% |
| Post radiofrequency catheter ablation | VIF | - | 0% |
| Post PCI / CABG | VIF | - | 0% |
| IVST (mm) | VIF | - | 0% |
| LVEDD (mm) | VIF | - | 0% |
| LVESD (mm) | VIF | - | 0% |
| LVMI (g/m<sup>2</sup>) | VIF | - | 0% |
| Left atrial diameter (mm) | VIF | - | 0% |
| BNP (pg/mL) | VIF | - | 1% |
| Creatinine (mg/dL) | VIF | - | 0% |
| TP (g/dL) | VIF | - | 0% |
| APTT (s) | VIF | - | 8% |

Abbreviations: PCI = percutaneous coronary intervention; CABG = coronary artery bypass grafting; IVST = Interventricular septal thickness; LVPWT = Left ventricular posterior wall thickness; LVEF = Left ventricular ejection fraction; LVEDD = Left ventricular end-diastolic diameter; LVESD = Left ventricular end-systolic diameter; LVMI = Left ventricular mass index.

**Supplemental Table 4.**
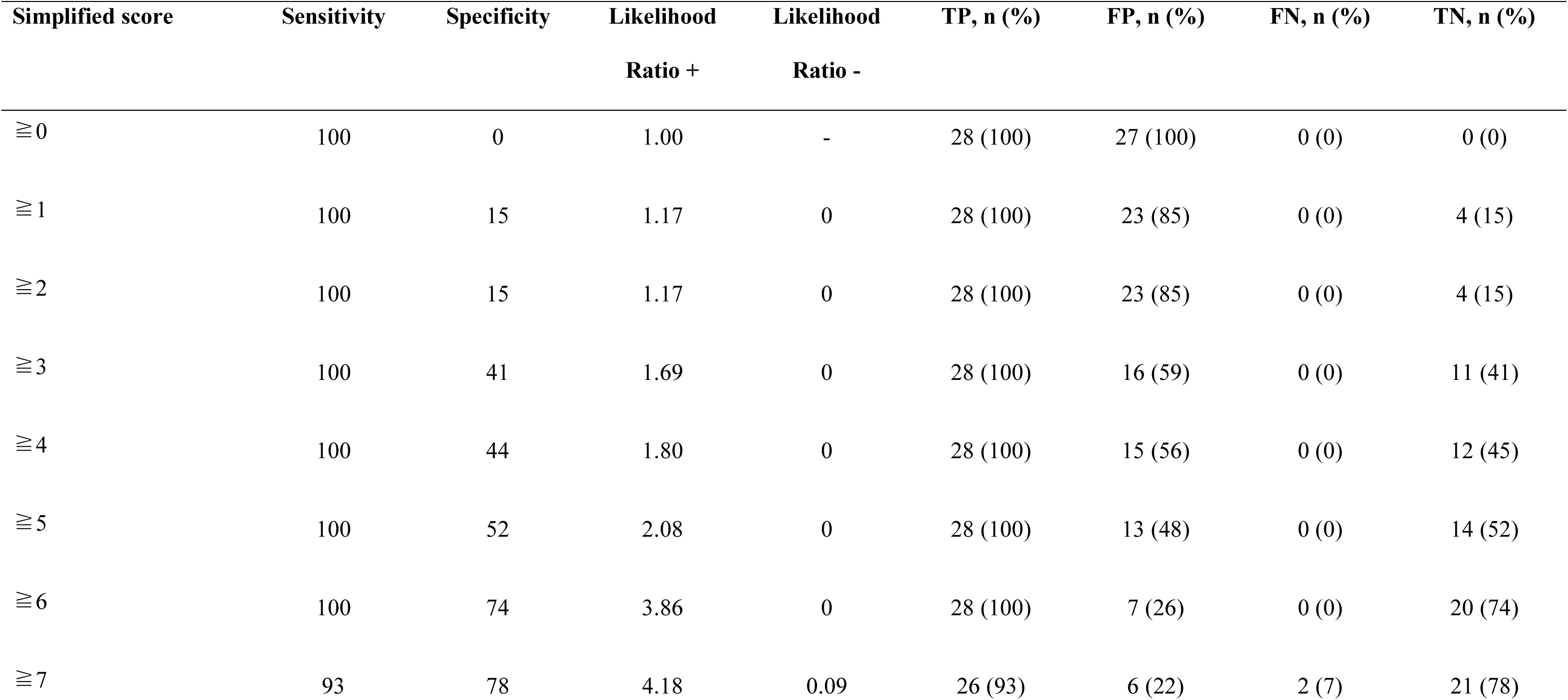
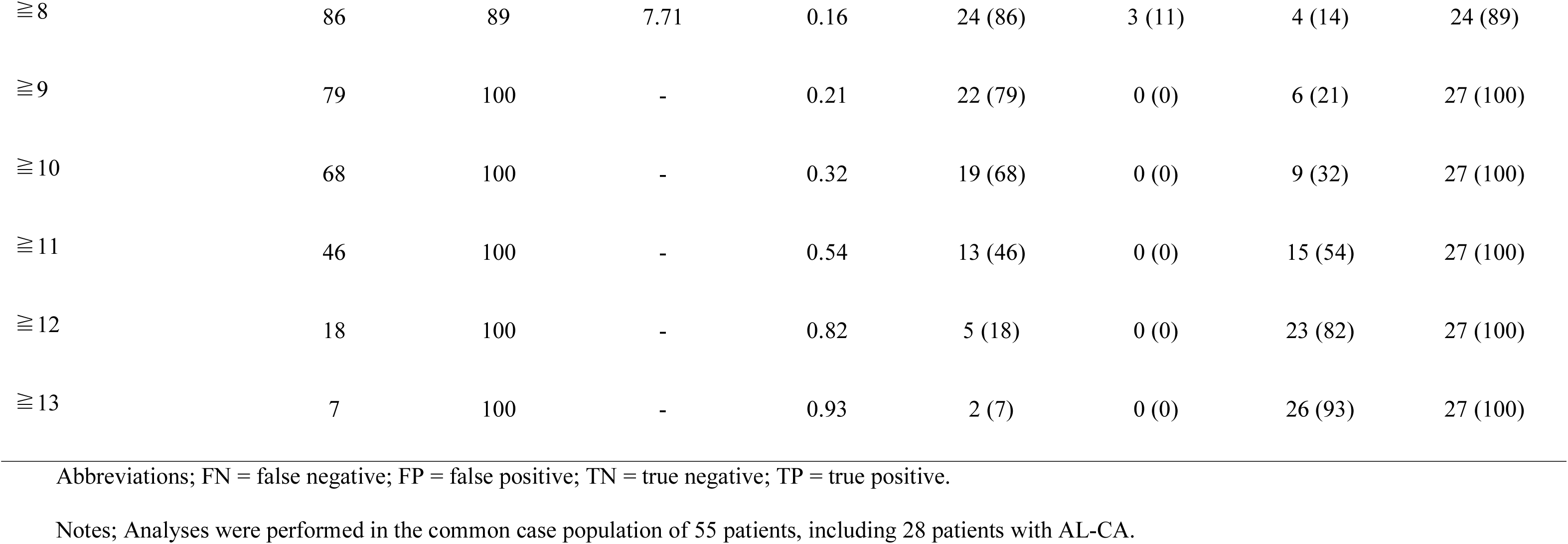
Diagnostic performance of the simplified score among patients with AL-CA or monoclonal protein–positive ATTRwt-CA.

| Simplified score | Sensitivity | Specificity | Likelihood<br>Ratio + | Likelihood<br>Ratio - | TP, n (%) | FP, n (%) | FN, n (%) | TN, n (%) |
| --- | --- | --- | --- | --- | --- | --- | --- | --- |
| ≧0 | 100 | 0 | 1.00 | - | 28 (100) | 27 (100) | 0 (0) | 0 (0) |
| ≧1 | 100 | 15 | 1.17 | 0 | 28 (100) | 23 (85) | 0 (0) | 4 (15) |
| ≧2 | 100 | 15 | 1.17 | 0 | 28 (100) | 23 (85) | 0 (0) | 4 (15) |
| ≧3 | 100 | 41 | 1.69 | 0 | 28 (100) | 16 (59) | 0 (0) | 11 (41) |
| ≧4 | 100 | 44 | 1.80 | 0 | 28 (100) | 15 (56) | 0 (0) | 12 (45) |
| ≧5 | 100 | 52 | 2.08 | 0 | 28 (100) | 13 (48) | 0 (0) | 14 (52) |
| ≧6 | 100 | 74 | 3.86 | 0 | 28 (100) | 7 (26) | 0 (0) | 20 (74) |
| ≧7 | 93 | 78 | 4.18 | 0.09 | 26 (93) | 6 (22) | 2 (7) | 21 (78) |
| $\geq 8$ | 86 | 89 | 7.71 | 0.16 | 24 (86) | 3 (11) | 4 (14) | 24 (89) |
| $\geq 9$ | 79 | 100 | - | 0.21 | 22 (79) | 0 (0) | 6 (21) | 27 (100) |
| $\geq 10$ | 68 | 100 | - | 0.32 | 19 (68) | 0 (0) | 9 (32) | 27 (100) |
| $\geq 11$ | 46 | 100 | - | 0.54 | 13 (46) | 0 (0) | 15 (54) | 27 (100) |
| $\geq 12$ | 18 | 100 | - | 0.82 | 5 (18) | 0 (0) | 23 (82) | 27 (100) |
| $\geq 13$ | 7 | 100 | - | 0.93 | 2 (7) | 0 (0) | 26 (93) | 27 (100) |

Abbreviations; FN = false negative; FP = false positive; TN = true negative; TP = true positive.
Notes; Analyses were performed in the common case population of 55 patients, including 28 patients with AL-CA.

**Supplemental Table 5.** Discriminative performance and pairwise comparison of FLC abnormality, immunofixation positivity, and the simplified score for identifying AL-CA among patients with AL-CA or ATTRwt-CA accompanied by monoclonal gammopathy Panel A. Discriminative performance of each model, Panel B. Pairwise comparison of model performance.

**Panel A. Discriminative performance of each model**
| Model | AUC | 95% CI |
| --- | --- | --- |
| FLC model | 0.54 | 0.43–0.65 |
| Immunofixation model | 0.69 | 0.57–0.82 |
| Score model (Simplified score $\geq 8$) | 0.90 | 0.82–0.99 |

**Panel B. Pairwise comparison of model performance**
| Reference model | New model | ΔAUC (95% CI) | DeLong p-value | Continuous NRI | p-value | IDI | p-value |
| --- | --- | --- | --- | --- | --- | --- | --- |
| FLC model | Immunofixation model | 0.15 (–0.012–0.32) | 0.068 | 0.77 | 0.003 | 0.14 | 0.007 |
| FLC model | Score model | 0.37 (0.22–0.51) | <0.001 | 1.62 | <0.001 | 0.64 | <0.001 |
| Immunofixation model | Score model | 0.21 (0.055–0.37) | 0.008 | 1.62 | <0.001 | 0.51 | <0.001 |

Abbreviations: AL, immunoglobulin light-chain amyloidosis; AUC, area under the receiver operating characteristic curve; CI, confidence interval; FLC, free light-chain ratio abnormality; IDI, integrated discrimination improvement; NRI, net reclassification improvement.
Notes; Analyses were performed in the common complete-case population of 52 patients, including 26 patients with AL-CA. AUC confidence intervals and pairwise differences in AUC were estimated using DeLong's method. ΔAUC was calculated as the AUC of the new model minus that of the reference model. NRI was calculated as continuous, category-free NRI. NRI and IDI p values were based on asymptotic Wald tests.

## References

1. Merlini G, Bellotti V. Molecular mechanisms of amyloidosis. N Engl J Med 2003;349:583–596.

2. Griffin JM, Maurer MS. Transthyretin cardiac amyloidosis: A treatable form of heart failure with a preserved ejection fraction. Trends Cardiovasc Med 2021;31:59–66.

3. Kwok CS, Moody WE. The importance of pathways to facilitate early diagnosis and treatment of patients with cardiac amyloidosis. Ther Adv Cardiovasc Dis 2023;17:17539447231216318.

4. Westin O, Butt JH, Gustafsson F, Schou M, Salomo M, Køber L, Maurer M, Fosbøl EL. Two Decades of Cardiac Amyloidosis: A Danish Nationwide Study. JACC CardioOncol 2021;3:522–533.

5. Kastritis E, Palladini G, Minnema MC, Wechalekar AD, Jaccard A, Lee HC, Sanchorawala V, Gibbs S, Mollee P, Venner CP, et al. Daratumumab-Based Treatment for Immunoglobulin Light-Chain Amyloidosis. N Engl J Med 2021;385:46–58.

6. Maurer MS, Schwartz JH, Gundapaneni B, Elliott PM, Merlini G, Waddington-Cruz M, Kristen AV, Grogan M, Witteles R, Damy T, et al. Tafamidis Treatment for Patients with Transthyretin Amyloid Cardiomyopathy. N Engl J Med 2018;379:1007–1016.

7. Fontana M, Berk JL, Gillmore JD, Witteles RM, Grogan M, Drachman B, Damy T, Garcia-Pavia P, Taubel J, Solomon SD, et al. Vutrisiran in Patients with Transthyretin Amyloidosis with Cardiomyopathy. N Engl J Med 2025;392:33–44.

8. Oubari S, Naser E, Papathanasiou M, Luedike P, Hagenacker T, Thimm A, Rischpler C, Kessler L, Kimmich C, Hegenbart U, et al. Impact of time to diagnosis on Mayo stages, treatment outcome, and survival in patients with AL amyloidosis and cardiac involvement. Eur J Haematol 2021;107:449–457.

9. Wechalekar AD, Fontana M, Quarta CC, Liedtke M. AL Amyloidosis for Cardiologists: Awareness, Diagnosis, and Future Prospects: JACC: CardioOncology State-of-the-Art Review. JACC CardioOncol 2022;4:427–441.

10. Oerlemans M, Rutten KHG, Minnema MC, Raymakers RAP, Asselbergs FW, de Jonge N. Cardiac amyloidosis: the need for early diagnosis. Neth Heart J 2019;27:525–536.

11. Morikawa K, Izumiya Y, Takashio S, Kawano Y, Oguni T, Kuyama N, Oike F, Yamamoto M, Tabata N, Ishii M, et al. Early experience with daratumumab-containing regimens in patients with light-chain cardiac amyloidosis. J Cardiol 2025;85:440–446.

12. Lyons G, Thompson J, Lousada I, Catini J, Manwani R, Maurer MS. Diagnostic pathways, cardiac manifestations and outcomes in light chain amyloidosis: analysis of a US claims database. Open Heart 2025;12:e003124.

13. Naito T, Nakamura K, Abe Y, Watanabe H, Sakuragi S, Katayama Y, Kihara H, Okizaki A, Kawai Y, Yoshikawa M, et al. Prevalence of transthyretin amyloidosis among heart failure patients with preserved ejection fraction in Japan. ESC Heart Fail 2023;10:1896–1906.

14. Kittleson MM, Ruberg FL, Ambardekar AV, Brannagan TH, Cheng RK, Clarke JO, Dember LM, Frantz JG, Hershberger RE, Maurer MS, et al. 2023 ACC Expert Consensus Decision Pathway on Comprehensive Multidisciplinary Care for the Patient With Cardiac Amyloidosis: A Report of the American College of Cardiology Solution Set Oversight Committee. J Am Coll Cardiol 2023;81:1076–1126.

15. Dorbala S, Ando Y, Bokhari S, Dispenzieri A, Falk RH, Ferrari VA, Fontana M, Gheysens O, Gillmore JD, Glaudemans AWJM, et al. ASNC/AHA/ASE/EANM/HFSA/ISA/SCMR/SNMMI Expert Consensus Recommendations for Multimodality Imaging in Cardiac Amyloidosis: Part 2 of 2-Diagnostic Criteria and Appropriate Utilization. Circ Cardiovasc Imaging 2021;14:e000030.

16. Nishihara T, Kawano Y, Tasaki M, Naito A, Yuki H, Nagayoshi Y, Takashio S, Nishimura N, Ueda M, Tsujita K, et al. Concurrent Immunoglobulin Light Chain and Transthyretin Cardiac Amyloidosis Patient Treated With Daratumumab and Tafamidis: A Case Report. EJHaem 2025;6:e70122.

17. Rajkumar SV, Kyle RA, Therneau TM, Melton LJ 3rd, Bradwell AR, Clark RJ, Larson DR, Plevak MF, Dispenzieri A, Katzmann JA. Serum free light chain ratio is an independent risk factor for progression in monoclonal gammopathy of undetermined significance. Blood 2005;106:812–817.

18. Collins GS, Reitsma JB, Altman DG, Moons KG. Transparent reporting of a multivariable prediction model for individual prognosis or diagnosis (TRIPOD): the TRIPOD statement. BMJ 2015;350:g7594.

19. Pinney JH, Whelan CJ, Petrie A, Dungu J, Banypersad SM, Sattianayagam P, Wechalekar A, Gibbs SD, Venner CP, Wassef N, et al. Senile systemic amyloidosis: clinical features at presentation and outcome. J Am Heart Assoc 2013;2:e000098.

20. White IR, Royston P, Wood AM. Multiple imputation using chained equations: Issues and guidance for practice. Stat Med 2011;30:377–399.

21. Dreyer RP, Raparelli V, Tsang SW, D’Onofrio G, Lorenze N, Xie CF, Geda M, Pilote L, Murphy TE. Development and Validation of a Risk Prediction Model for 1-Year Readmission Among Young Adults Hospitalized for Acute Myocardial Infarction. J Am Heart Assoc 2021;10:e021047.

22. Vickers AJ, Elkin EB. Decision curve analysis: a novel method for evaluating prediction models. Med Decis Making 2006;26:565–574.

23. Boldrini M, Cappelli F, Chacko L, Restrepo-Cordoba MA, Lopez-Sainz A, Giannoni A, Aimo A, Baggiano A, Martinez-Naharro A, Whelan C, et al. Multiparametric Echocardiography Scores for the Diagnosis of Cardiac Amyloidosis. JACC Cardiovasc Imaging 2020;13:909–920.

24. Dungu JN, Valencia O, Pinney JH, Gibbs SD, Rowczenio D, Gilbertson J, Lachmann HJ, Wechalekar AD, Gillmore JD, Whelan CJ, et al. CMR-based differentiation of AL and ATTR cardiac amyloidosis. JACC Cardiovasc Imaging 2014;7:133–142.

25. Poterucha TJ, Elias P, Bokhari S, Einstein AJ, DeLuca A, Kinkhabwala M, Johnson LL, Flaherty KR, Saith SE, Griffin JM, et al. Diagnosing Transthyretin Cardiac Amyloidosis by Technetium Tc 99m Pyrophosphate: A Test in Evolution. JACC Cardiovasc Imaging 2021;14:1221–1231.

26. Schafer EB, Tushak Z, Trankle CR, Rao K, Cartagena LC, Shah KB. False- Positive (99m)Technetium-Pyrophosphate Scintigraphy in Two Patients With Hypertrophic Cardiomyopathy. Circ Heart Fail 2021;14:e007558.

27. Izumiya Y, Kubo T, Endo J, Takashio S, Minamisawa M, Hamada J, Ishii T, Abe H, Konishi H, Tsujita K. Transthyretin amyloid cardiomyopathy: Literature review and red-flag symptom clusters for each cardiology specialty. ESC Heart Fail 2025;12:955–967.

28. Shafqat A, Elmaleh H, Mushtaq A, Firdous Z, Ashruf O, Mukhopadhyay D, Ahmad M, Ahmad M, Raza S, Anwer F. Renal AL Amyloidosis: Updates on Diagnosis, Staging, and Management. J Clin Med 2024;13:1744.

29. Patel RK, Fontana M, Ruberg FL. Cardiac Amyloidosis: Multimodal Imaging of Disease Activity and Response to Treatment. Circ Cardiovasc Imaging 2021;14:e009025.

30. Argirò A, Zampieri M, Mazzoni C, Fumagalli C, Baccini M, Mattei A, Cipriani A, De Michieli L, Porcari A, Sinagra G, et al. Progression and prognostic significance of electrocardiographic findings in patients with cardiac amyloidosis. ESC Heart Fail 2025;12:809–818.

31. Brenner DA, Jain M, Pimentel DR, Wang B, Connors LH, Skinner M, Apstein CS, Liao R. Human amyloidogenic light chains directly impair cardiomyocyte function through an increase in cellular oxidant stress. Circ Res 2004;94:1008–1010.

